# Two distinct immunopathological profiles in autopsy lungs of COVID-19

**DOI:** 10.1101/2020.06.17.20133637

**Authors:** Ronny Nienhold, Yari Ciani, Viktor H. Koelzer, Alexandar Tzankov, Jasmin D. Haslbauer, Thomas Menter, Nathalie Schwab, Maurice Henkel, Angela Frank, Veronika Zsikla, Niels Willi, Werner Kempf, Thomas Hoyler, Mattia Barbareschi, Holger Moch, Markus Tolnay, Gieri Cathomas, Francesca Demichelis, Tobias Junt, Kirsten D. Mertz

## Abstract

Coronavirus Disease 19 (COVID-19) is a respiratory disease caused by severe acute respiratory syndrome coronavirus 2 (SARS-CoV-2), which has grown to a worldwide pandemic with substantial mortality. Immune mediated damage has been proposed as a pathogenic factor, but immune responses in lungs of COVID-19 patients remain poorly characterized. Therefore we conducted transcriptomic, histologic and cellular profiling of *post mortem* COVID-19 (n=34 tissues from 16 patients) and normal lung tissues (n=9 tissues from 6 patients). Two distinct immunopathological reaction patterns of lethal COVID-19 were identified. One pattern showed high local expression of interferon stimulated genes (ISG^high^) and cytokines, high viral loads and limited pulmonary damage, the other pattern showed severely damaged lungs, low ISGs (ISG^low^), low viral loads and abundant infiltrating activated CD8+ T cells and macrophages. ISG^high^ patients died significantly earlier after hospitalization than ISG^low^ patients. Our study may point to distinct stages of progression of COVID-19 lung disease and highlights the need for peripheral blood biomarkers that inform about patient lung status and guide treatment.

## Introduction

COVID-19 is a pandemic respiratory disease with 2-3% lethality and a particularly severe course in males, patients with cardiovascular comorbidities and in the elderly^1,2^. Lymphopenia, high levels of pro-inflammatory cytokines in the circulation^3^, and phenotypic changes of pro-inflammatory macrophages in bronchoalveolar lavages^4^ in severe patients have led to the notion that the immune response against the causative virus SARS-CoV-2 may contribute to devastating end organ damage^5^. Since patients with severe COVID-19 may develop acute respiratory distress syndrome (ARDS) and many patients die from respiratory failure with diffuse alveolar damage^6^, it is critical to understand the immunological profiles in the lungs of these patients.

To better understand the molecular and cellular underpinnings of COVID-19 lung disease, we used histologic and transcriptional analyses of *post mortem* lung tissues in a cohort of patients where the cause of death was respiratory failure. We detected two distinct immunological and cellular profiles in the lungs of these patients, defined by their differential expression of interferon stimulated genes (ISGs) and immune infiltration patterns, which we termed ISG^high^ and ISG^low^. ISG subgroups strongly differed in regards to the characteristics and the extent of pulmonary damage, pulmonary viral loads, disease course and time to death from hospitalization. These data highlight two distinct patterns of immune pathology of pulmonary COVID-19 and may give insight into the natural progression of COVID-19 in lungs.

## Results

### Two patterns of gene expression in COVID-19 autopsy lungs

Here we analyzed 34 *post mortem* lung samples from 16 deceased COVID-19 patients and 9 *post mortem* lung samples from 6 patients, who died from non-infectious causes (**Table 1**). The primary cause of death in all patients of this cohort was respiratory failure, sometimes multi-organ failure including failure of the respiratory system. Since lung samples from the same patients did not always appear morphologically uniform, all lung specimens were subjected to differential gene expression analysis based on a commercially available targeted next generation sequencing (NGS) assay (OIRRA) designed for quantification of immune cell and inflammatory transcripts (**Supplementary Table 2**). Among the 398 genes investigated, we identified 68 up-regulated and 30 down-regulated genes in COVID-19 infected lungs compared to normal tissue (**Figure 1a,b**; **Supplementary Table 3**), and a PCA analysis showed segregation of COVID-19 patients in two well-defined clusters that showed distinct association with viral load (**Figure 1a,c**).

**Table 1a.** Clinical Data, COVID-19 cohort (16 patients)

| Characteristics | N or x | % or range |
| --- | --- | --- |
| Time between symptoms and death | 7,4 d | 0 - 20 d |
| Hospitalisation | 5,6 d | 0 - 13 d |
| Postmortem interval | 28,4 h | 11 - 67 h |
| Age | 75 y | 53 - 96 y |
| Sex | M 13 : F 3 |  |
| Comorbidities |  |  |
| Hypertension | 16 | 100 % |
| Cardiovascular disease | 11 | 68 % |
| (Pre-)Adipositas | 12 | 75 % |
| Diabetes | 6 | 37,5 % |
| Initial clinical presentation |  |  |
| Cough | 13 | 81,25 % |
| Fever | 12 | 75 % |
| Dyspnea | 6 | 37,5 % |
| Renal failure | 6 | 37,5 % |
| Laboratory results |  |  |
| Interleukin-6 (IL6) | 5774,72 ng/l | 159,00 - 35.152,00 ng/l |
| C-reactive protein (CRP) | 216,36 mg/l | 36,00 - 512,00 mg/l |
| Ferritin | 18.037,71 µg/l | 1.025,00 - 228.225,00 µg/l |
| Procalcitonin (PCT) | 1.996,92 µg/l | 0,47 - 5.300,00 µg/l |
| Lactate dehydrogenase (LDH) | 870,42 U/l | 256,00 - 5.267,00 U/l |
| Troponin T (cTnT) | 54,65 ng/l | 2,07 - 126,00 ng/l |
| Treatment |  |  |
| Hydroxychloroquine | 10 | 62,5 % |
| Lopinavir/Ritonavir | 5 | 31,25 % |
| Antibiotics | 12 | 75 % |
| ACTEMRA (Tocilizumab) | 5 | 31,25 % |

Listed laboratory results correspond to the highest (LDH, cTnT), latest (IL6, ferritin, PCT) or last value before administration of ACTEMRA/Tocilizumab (CRP).

**Table 1b.** Clinical Data, Control cohort (6 patients)

| Characteristics | N or x | % or range |
| --- | --- | --- |
| Postmortem interval | 31,4 h | 25 - 46 h |
| Age | 81,3 y | 63 - 104 y |
| Sex | M 5 : F 1 |  |

**Table 1c.** Clinical Data, Cohort of patients with other infections (4 patients)

| Characteristics | N or x | % or range |
| --- | --- | --- |
| Postmortem interval | 31,5 h | 12 - 79 h |
| Age | 79,5 y | 58 - 81 y |
| Sex | M 2 : F 2 |  |

**Figure 1.**
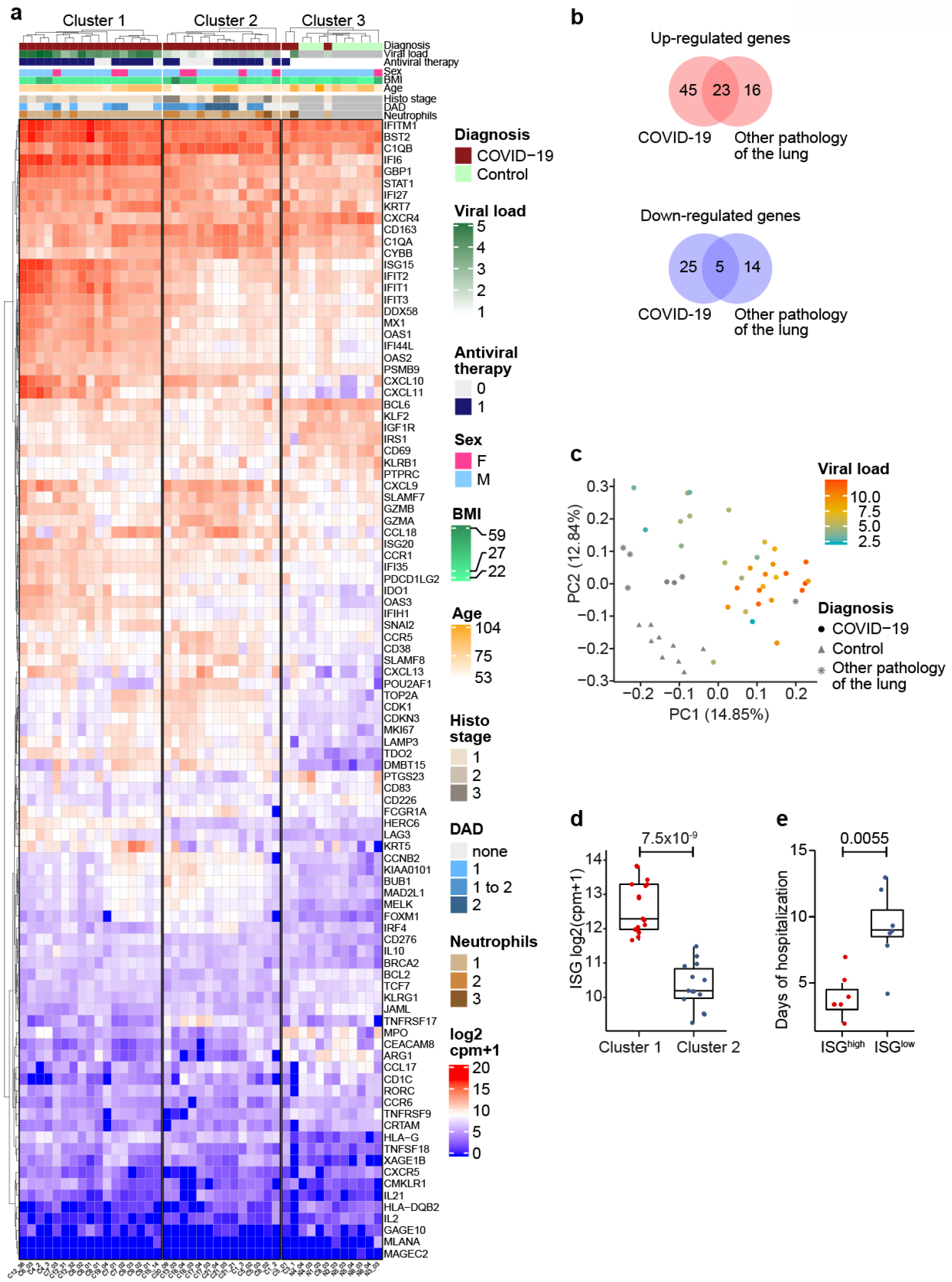
ISG^high^ and ISG^low^ are two gene expression profiles in COVID-19 autopsy lungs. (**a**) Heatmap showing K-means clustering of COVID-19 and normal lung samples based on expression levels of deregulated genes in COVID-19 versus normal lungs. (**b**) Comparison of up- and down-regulated genes in lung samples from COVID-19 patients, normal lung samples and samples from other infectious lung pathologies. (**c**) Principal component analysis (PCA) of COVID-19 and non-COVID-19 lung samples reveals segregation in two distinct groups based on diagnosis and viral load. (**d**) ISG signature expression in clusters 1 and 2 of COVID-19 lungs defines two profiles of COVID-19 autopsy lungs termed ISG^high^ and ISG^low^. Study patients with unambiguous sample segregation in either Cluster 1 or 2 were assigned the corresponding ISG activation label ISG^high^ and ISG^low^, respectively. (**e**) Hospitalization time in ISG^high^ patients versus ISG^low^ patients. ISG^high^ samples, red; ISG^low^ samples, blue

Using a consensus of 30 different indices^7^ we identified three groups of samples defined by distinct expression levels of the deregulated genes by K-means clustering (**Figure 1a**). Clusters 1 (50% of samples) and 2 (41%) contained COVID-19 samples while cluster 3 contained all normal lung samples as well as three COVID-19 samples (9%). To understand why the majority of COVID-19 lung tissues segregated into defined clusters 1 or 2, we undertook a gene ontology analysis. We identified ISGs as a key upregulated pathway in COVID-19 autopsy lungs (**Table 2**), which was differentially represented in cluster 1 and 2, respectively (**Figure 1d**). Identification of an ISG^high^ cluster (Cluster 1, ISG^high^) was surprising, since SARS-CoV-2 was recently proposed to lead to limited ISG induction, yet only in comparison to other respiratory viruses^8^. Our data suggest that autopsy lungs of COVID-19 patients, who died from respiratory failure, showed two different gene expression patterns with different levels of ISG activation (ISG^high^ and ISG^low^).

**Table 2.** Gene ontology enrichment analysis of genes upregulated in COVID-19 samples.

| **ID** | **Description** | **GeneRatio** | **BgRatio** | **pvalue** | **p.adjust** |
| --- | --- | --- | --- | --- | --- |
| GO:0034340 | response to type I interferon | 17/66 | 30/379 | 3,70E-07 | 0,000158 |
| GO:0060337 | type I interferon signaling pathway | 17/66 | 30/379 | 3,70E-07 | 0,000158 |
| GO:0071357 | cellular response to type I interferon | 17/66 | 30/379 | 3,70E-07 | 0,000158 |
| GO:0051607 | defense response to virus | 21/66 | 44/379 | 5,53E-07 | 0,000177 |
| GO:0098542 | defense response to other organism | 25/66 | 71/379 | 3,85E-05 | 0,009888 |
| GO:0009615 | response to virus | 21/66 | 57/379 | 9,79E-05 | 0,020960 |
| GO:0045069 | regulation of viral genome replication | 10/66 | 18/379 | 1,87E-04 | 0,034381 |
| GO:1903900 | regulation of viral life cycle | 11/66 | 22/379 | 2,95E-04 | 0,042931 |
| GO:0045071 | negative regulation of viral genome replication | 8/66 | 13/379 | 3,57E-04 | 0,042931 |
| GO:1903901 | negative regulation of viral life cycle | 8/66 | 13/379 | 3,57E-04 | 0,042931 |
| GO:0048525 | negative regulation of viral process | 9/66 | 16/379 | 3,68E-04 | 0,042931 |
| GO:0051603 | proteolysis involved in cellular protein catabolic process | 7/66 | 11/379 | 6,71E-04 | 0,071789 |
| GO:0050792 | regulation of viral process | 12/66 | 28/379 | 9,17E-04 | 0,088399 |
| GO:0019079 | viral genome replication | 10/66 | 21/379 | 9,64E-04 | 0,088399 |
| GO:0043901 | negative regulation of multi-organism process | 9/66 | 18/379 | 1,16E-03 | 0,099123 |

### Clinical differences of COVID-19 patients with an ISG^high^ versus ISG^low^ lung profile

Patients, whose lung samples all segregated in the ISG^high^ or in the ISG^low^ subgroups, were called ISG^high^ and ISG^low^ patients. To investigate whether there were clinical differences between these two patient groups, we compared their clinical and epidemiological information. The majority of COVID-19 patients in our cohort (81%) were male and the average body mass index (BMI) was 31.4. kg/m^2^. Patient level analysis revealed no correlation of sex or BMI with the ISG patterns (**Figure 1a**). All patients in our cohort died from respiratory failure or multi-organ failure including failure of the respiratory system, independent of ISG subgrouping. When we analysed comorbidities and autopsy findings, we found that 5 out of 7 (71%) ISG^low^ patients, but none of the ISG^high^ patients had an autoptic finding of a thromboembolic event in the lungs and/or disseminated intravascular dissemination (DIC) indicating an abnormally activated blood coagulation (hypercoagulability) exclusively in ISG^low^ patients. COVID-19 associated coagulopathy was unlikely to contribute to rapid exacerbation of pulmonary COVID-19 since this diagnostic criterion was only observed in patients displaying the ISG^low^ phenotype. Most notably, when we compared the disease course and hospitalization time between the patients with different ISG lung profiles, we found a significantly longer hospitalization time for ISG^low^ patients from admission to death compared to ISG^high^ patients (**Figure 1e**). Of note, we did not find differences in the anamnestic onset of disease symptoms and time from positive testing for COVID-19 infection by nasopharyngeal swab to hospitalization or death between ISG^high^ and ISG^low^ patients. To exclude bacterial and viral superinfections as a confounder of clinical course, we performed whole genome sequencing on all samples to detect bacterial and/or viral DNA. Bacterial superinfections were found in three lung tissue samples, in 3/16 COVID-19 patients, that were equally distributed among the different groups (**Figure 2a-e**). Based on the limited number of samples with evidence of bacterial superinfection, there was no correlation with clinical subgroups or the duration of the disease.

**Figure 2.**
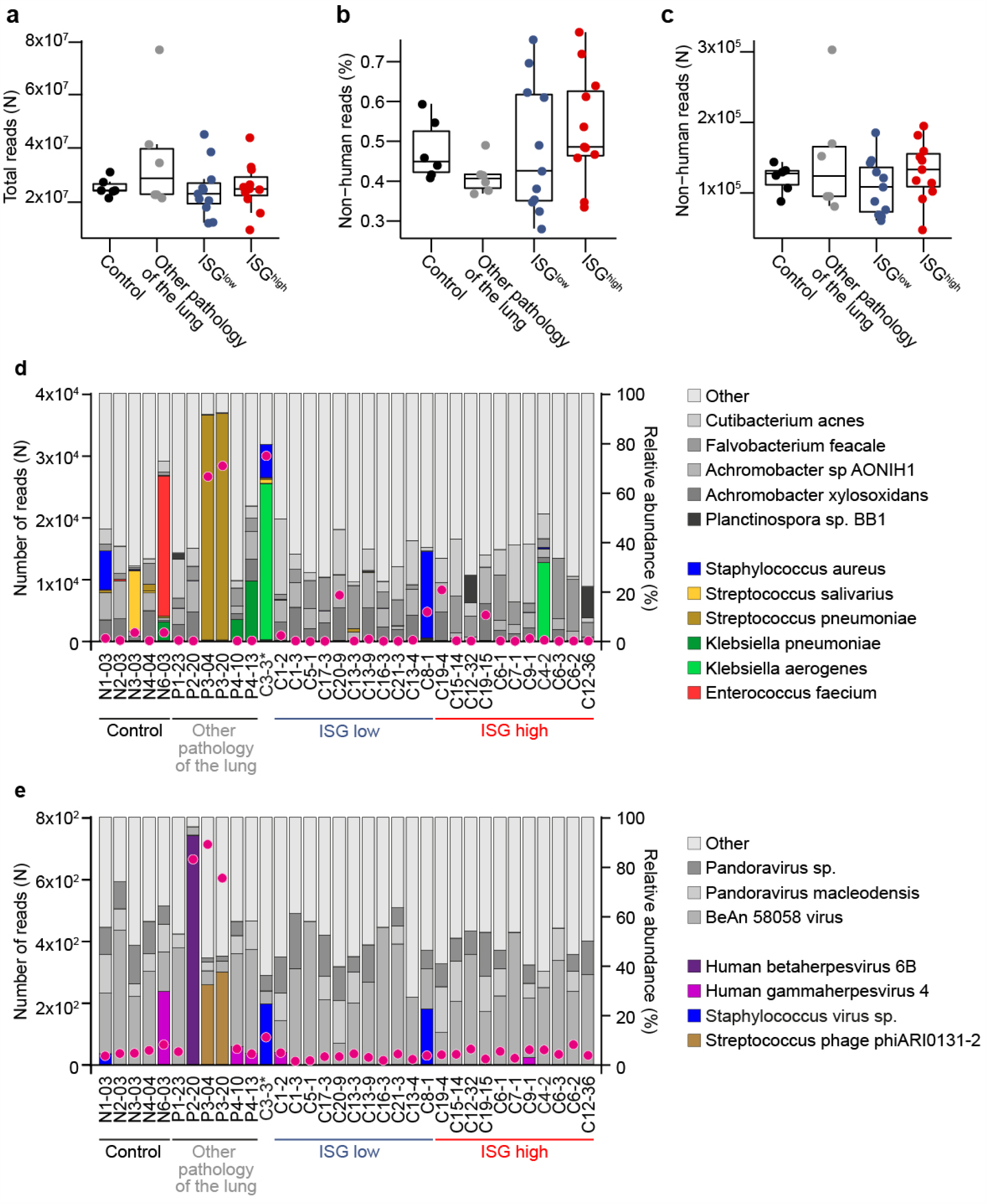
Co-infections in COVID-19 lungs identified by WGS metagenomics. No differences in co-infections in ISG^high^ and ISG^low^ COVID-19 lungs identified by WGS metagenomics. (**a**) Total number of reads generated for each sample. (**b**) Percentage of reads and (**c**) absolute numbers of reads not mapping to the human genome (GRCh37 hg19). (**d**) Bacterial and (**e**) viral co-infections across lung samples, WGS metagenomic analysis. Purple dots, numbers of reads sufficient for identification of non-human species. Samples are ordered by increasing SARS-CoV-2 viral load in both the ISG^low^ and the ISG^high^ group. Stacked bars, relative abundance of the most common species. Grey bars represent frequent species, colored bars show pathogenic species. *One COVID-19 patient (C3) clustered in the normal control group. ISG^high^ samples, red; ISG^low^ samples, blue

Taken together, expression of the ISG^high^ profile in COVID-19 lungs is associated with accelerated disease course and early lethal outcome. The ISG^low^ profile is associated with coagulopathies and later lethal outcome. This points to two distinct clinical courses of fatal COVID-19 pulmonary disease. Since autopsy studies always focus on disease endpoints, our data do not allow us to draw direct conclusions about natural stages of disease progression in COVID-19 lungs. However, it is very suggestive that the ISG^high^ profile precedes the ISG^low^ profile, consistent with a longitudinal study in peripheral blood showing that ISG expression was high in early COVID-19 and declined later^9^.

### Immune microenvironment characteristics of the ISG^high^ and ISG^low^ COVID-19 lung profiles

In line with a recent study showing a correlation of ISG expression and viral load in nasopharyngeal swabs^10^, expression of ISGs was positively correlated with pulmonary viral load (**Figure 3a**), and immunohistochemical staining confirmed the presence of SARS-CoV-2 nucleocapsid protein in ISG^high^ lungs, mainly localized to pneumocytes (**Figure 3b**).

**Figure 3.**
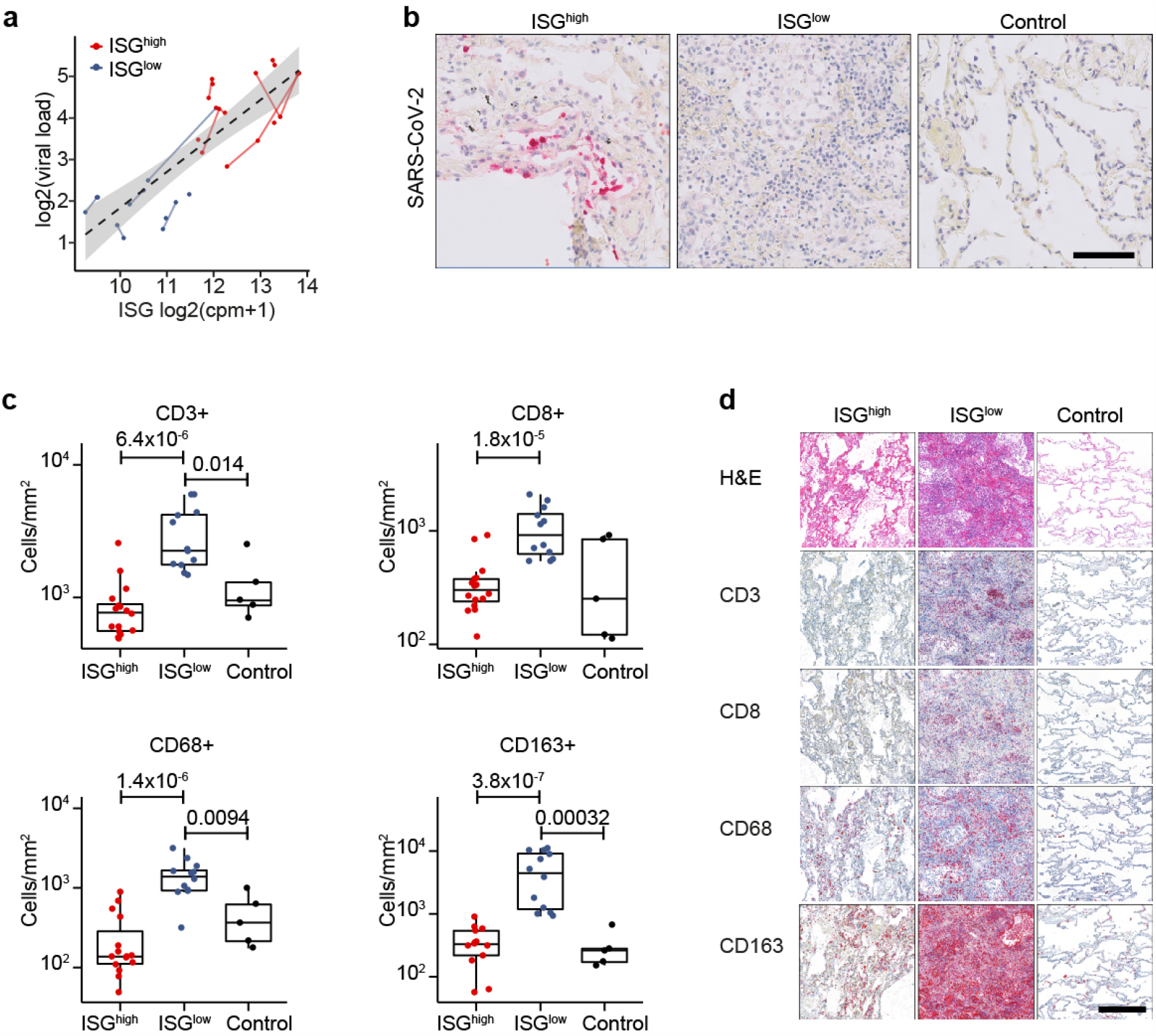
Virological and cellular characteristics of the ISG^high^ and ISG^low^ COVID-19 lung profiles. (**a**) Correlation of viral load and ISG expression in COVID-19 lungs. Solid lines, sample data from the same patient. Dotted line, regression for all samples. Grey, 95% CI (Pearson’s correlation=0.83, adjusted R-squared=0.68, p-value=1.66e-08). (**b**) Representative immunohistochemistry for SARS-CoV-2 on ISG^high^ and ISG^low^ COVID-19 lung samples and controls. Size bar 100 μm. (**c**) Frequencies of immune cells on ISG^high^ and ISG^low^ COVID-19 lung sections and controls. (**d**) Representative H&E stains and immunohistochemistry (CD3, CD8, CD68, CD163) of ISG^high^ and ISG^low^ COVID-19 lungs and controls, size bar 500 μm. ISG^high^ samples, red; ISG^low^ samples, blue

SARS-CoV-2 induces a strong antiviral immune response. Therefore we analysed frequencies of specific immune cells in lungs by computational image analysis. T cells (CD3+) of the CD4+ and CD8+ lineages, B cells (CD20+), and macrophages (CD68+) were selectively enriched in lung tissues from ISG^low^ patients (**Figure 3c,d**; **Figure 4a,b**). A strong enrichment for CD68+ and CD163+ monocytes in lung tissue was observed with spatial correlation of stains for both markers indicating co-expression. Since circulating monocytes in COVID-19 patients co-express CD68 and CD163, it was not surprising that CD68 and CD163 expression in lungs followed a similar pattern^11^ (**Figure 3c,d**). Surprisingly, CD123+ plasmacytoid dendritic cells (pDCs) did not show elevated frequencies in ISG^high^ lungs (**Figure 4a,b**), and the type 1 IFN (IFN-I) genes that we measured (IFNa17, IFNb1) were not higher expressed in ISG^high^ lungs with high viral load compared to ISG^low^ lungs (*data not shown*), potentially because SARS-CoV-2 inhibits IFN-I production^8^. Therefore our analysis did not allow us to identify the upstream trigger of ISGs in lungs.

**Figure 4.**
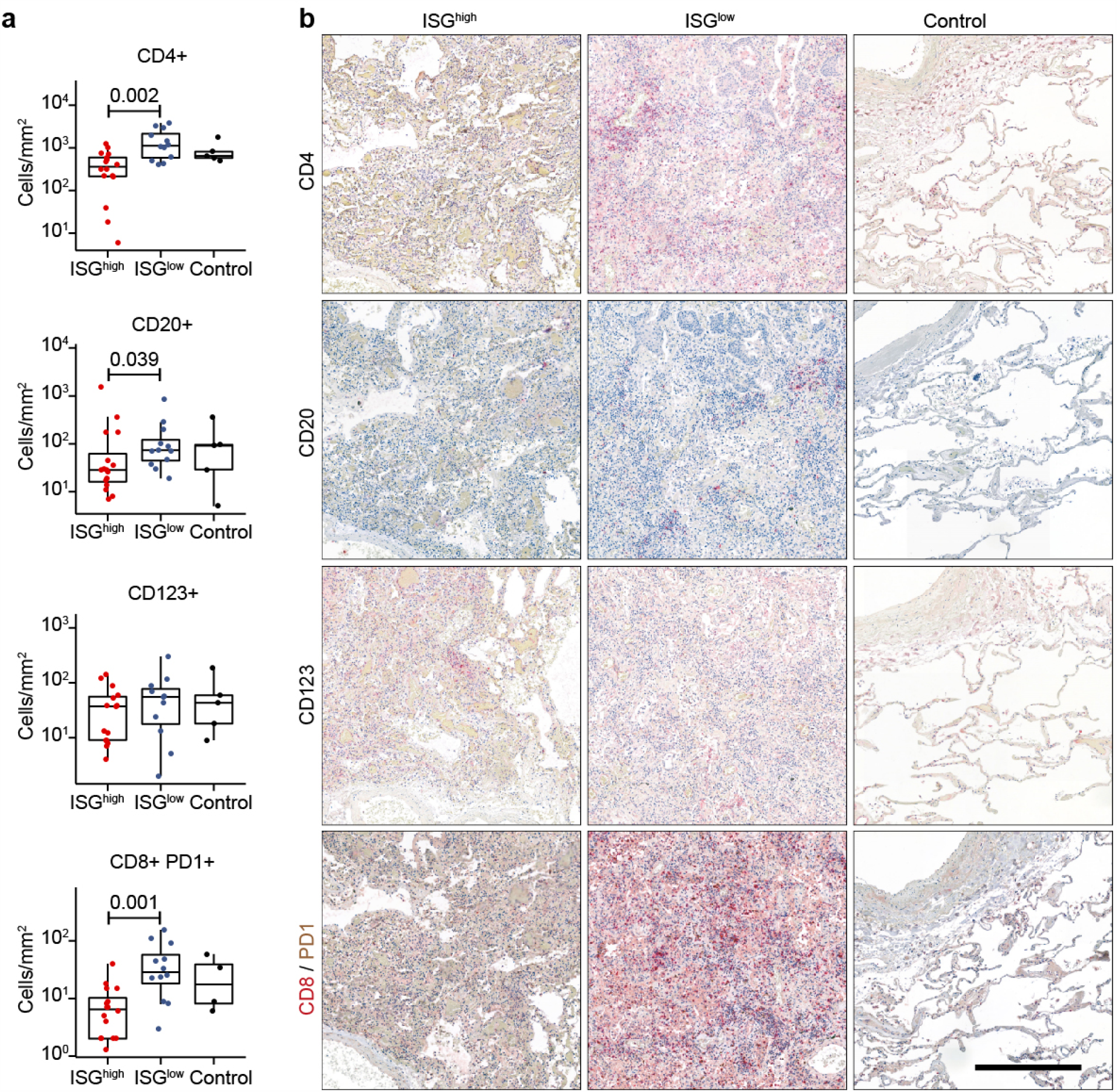
Immune cell infiltrates on COVID-19 lung sections. (**a**) Frequencies of immune cells on ISG^high^ and ISG^low^ COVID-19 lung sections and controls. (**b**) Representative immunohistochemistry (CD4, CD20, CD123, CD8/PD1) of ISG^high^ and ISG^low^ COVID-19 lungs and controls, size bar 500 μm. ISG^high^ samples, red; ISG^low^ samples, blue

A cytokine storm has been proposed to cause adverse outcome of COVID-19^12^. It has been suggested that peripheral monocytes do not contribute to it^13^, yet cytokines are highly expressed in bronchoalveolar lavages (BALs) of COVID-19 patients^4^. Therefore we investigated expression of a pro-inflammatory cytokine signature (TNF, IL1B, IL6, CCL2, IFNA17, IFNB1, CXCL9, CXCL10, CXCL11) in lung samples from lethal COVID-19. The proinflammatory gene signature was significantly enriched in the ISG^high^ subset (p=0.0061) (**Figure 5a**). Activated CD8+ T cells are essential for elimination of coronaviruses^14,15^. Therefore we defined and investigated an activated cytotoxic T cell signature (CD38, GZMA, GZMB, CCR5) and found that it was inversely correlated to viral counts, particularly in ISG^low^ cases (**Figure 5b**). This suggests that activated CD8+ T cells may indeed contribute to the elimination of SARS-CoV-2 in lungs.

**Figure 5.**
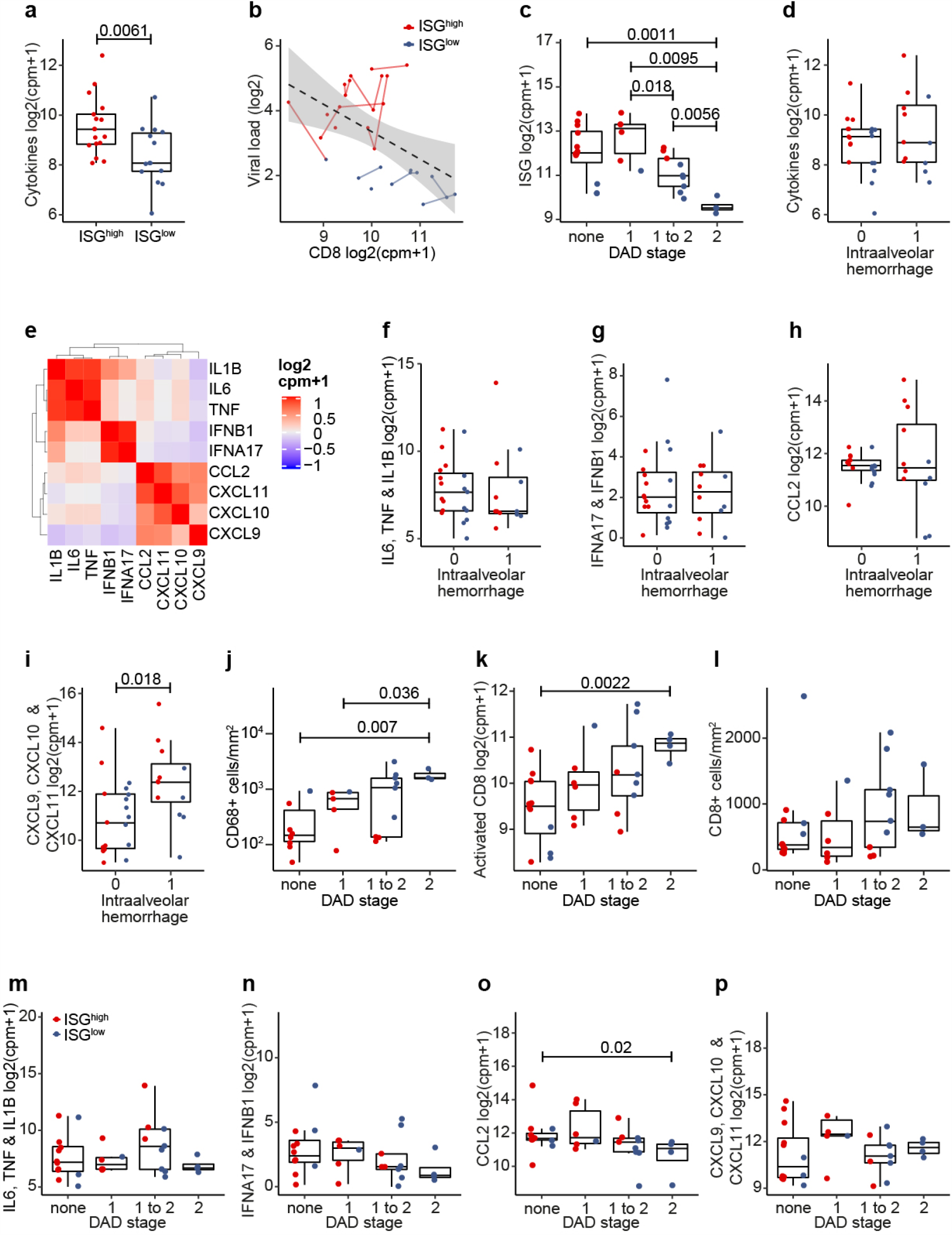
Correlation of ISG^high^ and ISG^low^ lung immunoprofiles with morphological changes. (**a**) Expression of a cytokine signature (TNF, IL-1B, IL6, IFNA17, IFNB1, CCL2, CXCL9, CXCL10, CXCL11) in ISG^high^ and ISG^low^ COVID-19 lung samples. This pro-inflammatory cytokine signature was significantly enriched in the ISG^high^ subset. (**b**) Inverse correlation of viral load and activated CD8+ T cell signature (CD38, GZMA, GZMB, CCR5). Solid lines, sample data from the same patient. Dotted line, regression for all the samples. Grey, 95% CI (Pearson’s correlation=-0.5, adjusted R-squared=0.22, p-value=0.005). (**c**) Association of DAD stage with ISG expression. (**d**) Association of the pro-inflammatory cytokine signature with intraalveolar hemorrhage (IAH). (**e**) Pearson’s correlation of pro-inflammatory cytokines in the cytokine signature indicates presence of co-regulated cytokines. (**f-i**) Association of cytokine signatures in ISG^high^ and ISG^low^ COVID-19 lung samples with IAH. Association of: (**f**) Median IL6, TNF, IL1B expression (**g**) Median IFNA17, IFNB1 expression (**h**) Median CCL2 expression (**i**) Median CXCL9/10/11 expression in ISG^high^ and ISG^low^ COVID-19 lung samples versus IAH. Only the CXCL9/10/11 sub-signature was positively associated with IAH. (**j**) Association of CD68+ macrophage infiltrates with DAD. (**k**) Association of DAD stage with activated CD8+ T cell signature, (**l**) with CD8+ T cell counts. (**m-p**) Association of cytokine signatures in ISG^high^ and ISG^low^ COVID-19 lung samples with DAD stage. Association of: (**m**) Median IL6, TNF, IL1B expression (**n**) Median IFNA17, IFNB1 expression (**o**) Median CCL2 expression (**p**) Median CXCL9/10/11 expression in ISG^high^ and ISG^low^ COVID-19 lung tissue with DAD stage. ISG^high^ samples, red; ISG^low^ samples, blue

Taken together these data show that COVID-19 autopsy lungs with an ISG^high^ profile show high virus titers, high local expression of innate cytokines and weak immune cell infiltration, while COVID-19 lungs with an ISG^low^ profile show low virus titers, lower expression of innate cytokines and strong immune cell infiltration. This pattern could indicate expression of the lung ISG^high^ profile at an earlier, innate disease stage, i.e. at a time when the virus is not yet controlled, and expression of the ISG^low^ profile at a later disease stage, i.e. after T cell priming.

### The ISG^high^ and ISG^low^ lung immunoprofiles correlate with morphological changes

To investigate the potential immunological causes for lung damage in COVID-19, we studied whether ISG profiles in COVID-19 *post mortem* lungs were associated with specific histomorphological features of fatal COVID-19. Diffuse alveolar damage (DAD) was mostly found in ISG^low^ patients (**Figure 5c**), but intra-alveolar hemorrhage (IAH) was not associated with lung ISG status.

As the cytokine storm was implicated in decline of COVID-19 patients, we analysed whether expression of the above defined pro-inflammatory cytokine signature was associated with IAH but this was not the case (**Figure 5d**). However, within this cytokine signature we identified co-regulated subgroups (IL1B/IL6/TNF, IFNB1/IFNA17, CCL2/CXCL9/CXCL19/CXCL11) (**Figure 5e**). Of these, the CXCL9/10/11 sub-signature was positively associated with IAH (**Figure 5f-i**). This is in line with observations that these chemokines compromise endothelial integrity via CXCR3^16^, and that CXCL10 is a key determinant of severe COVID-19^17^. Interestingly, basal levels of CXCL9 or CXCL10 are elevated in elderly, hypertensive and obese individuals, who were strongly represented in our autopsy cohort (**Table 1**) and are predisposed to severe COVID-19^18,19^.

It has been proposed that infiltrating monocytes and macrophages play a role in lung damage^4,20^. In support of this data, we found CD68+ macrophage infiltrates to be positively associated with DAD (**Figure 5j**). In addition, DAD was associated with the activated cytotoxic T cell signature (p=0.0022) (**Figure 5k**), yet not with the overall numbers of pulmonary CD8+ T-cells (**Figure 5l**). This raises the possibility that activated CD8+ T cells contribute to DAD as they eliminate virus from infected lungs. None of the above pulmonary cytokine sub-signatures, however, was positively associated with DAD (**Figure 5m-p**), suggesting that none of these cytokines drives lung pathology directly.

In summary, we did not find distinct features of lung damage in ISG^high^ patients, suggesting that extra-pulmonary factors may contribute to mortality in these patients. However, ISG^low^ patients show prominent DAD, associated with peri-alveolar foci of CD68+ macrophages and an activated T cell signature. Local expression of most cytokines did not correlate to lung damage, except for CXCL9/10/11, which correlated to IAH (p=0.018) (**Figure 5i**). Based on studies that associated serum CXCL10 levels with general disease severity^17^, it will be interesting to investigate whether serum levels of these cytokines predict a specific pattern of lung damage.

### The ISG^low^ lung profile shows signs of tissue regeneration and T cell exhaustion

Since ISG^low^ lung samples were derived from patients with a longer disease course, we investigated specific pathways of local immune regulation and tissue regeneration. ISG^low^ lung samples expressed elevated p53 and Ki67 (**Figure 1a, Figure 6a**), i.e. reactive markers indicating lung remodeling in DAD^21^.

**Figure 6.**
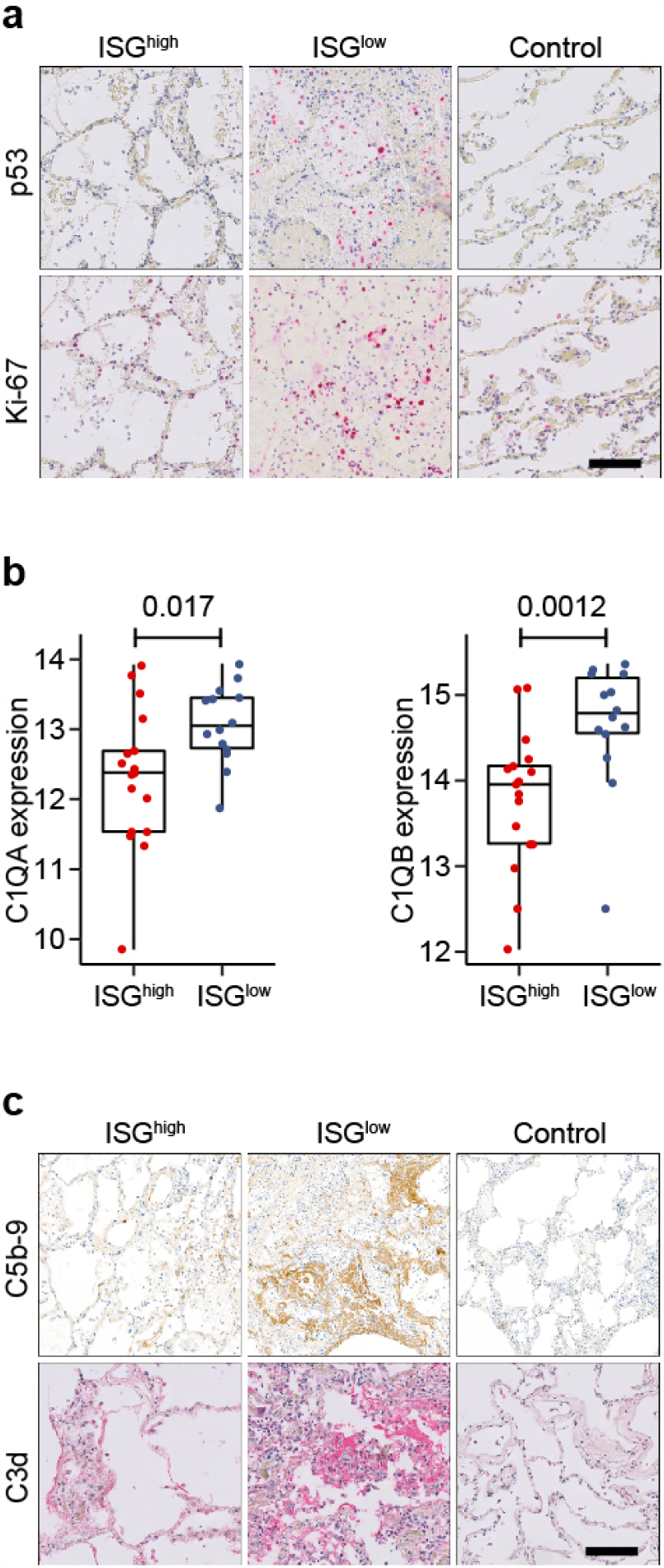
Molecular characteristics of the ISG^high^ and ISG^low^ COVID-19 lung profiles. (**a**) Representative immunohistochemistry for p53 and Ki67. Size bar 100 μm. (**b**) Expression of C1QA and C1QB in ISG^high^ and ISG^low^ lung samples. (**c**) Representative IHC stainings for complement activation products C5b-9 and C3d in ISG^high^, ISG^low^ COVID-19 and normal control lungs. Size bar 100 μm. ISG^high^ samples, red; ISG^low^ samples, blue

Since we found local upregulation of C1QA (p=0.017) and C1QB (p=0.0012) specifically in ISG^low^ lungs (**Figure 1a, Figure 6b**), we hypothesized that complement activation may further contribute to lung damage in these patients. Consistently, we found strong staining for C3d and C5b-9 complex deposition in lung tissue indicating complement activation in lungs of ISG^low^ patients (**Figure 6c**). Since C1Q also restrains antiviral CD8+ effector T cell responses^22^, it may contribute to the local regulation of effector T cells. In line with previous observations^23^, we found a higher frequency of CD8+PD1+ T-cells in the ISG^low^ subgroup (p=0.001), potentially indicative of T cell exhaustion (**Figure 4a,b**).

Overall our results identify two patterns of pulmonary COVID-19 disease that lead to death from respiratory failure. Patients of the ISG^high^ subgroup die early with high viral loads and high cytokine and ISG expression levels in lungs. Their lungs are morphologically relatively intact, and our data do not identify a uniform pathomechanism underlying lethal outcome, although some show CXCL9/10/11-associated IAH (**Figure 5i**). The distinct ISG^low^ group of patients dies later, with low viral loads in the lungs, low local expression of cytokines and ISGs, yet strong infiltration of pulmonary tissue by CD8+ T cells and macrophages, which both correlate to severity of DAD and local complement activation. Some of these patients show IAH in addition to DAD, and many of them suffer from coagulopathies. Altogether this patient group appears to suffer from severe pulmonary immunopathology. The design of our autopsy study does not allow to directly conclude that ISG^low^ lungs might have undergone a previous ISG^high^ phase, though circumstantial evidence about the general course of coronavirus infections may suggest this possibility.

## Discussion

Here we describe two immunopathological patterns in lungs of fatal COVID-19 patients based on ISG expression. The ISG^high^ pattern is observed in patients, who die early after hospitalization and is characterized by high viral load and high levels of pro-inflammatory cytokines, yet relatively intact lung morphology, while the ISG^low^ pattern is characterized by low viral load, massive lung damage, marked lung immune cell infiltrates and late death. Our findings are consistent with epidemiological data showing two peaks of mortality^24^, and another study of four COVID-19 autopsies, where one patient died early after hospital admission, with striking upregulation of pulmonary IL-1b/IL-6 in lungs and little lung damage, while three patients expressed low levels of pulmonary cytokines, massive DAD and delayed death^25^. Therefore our study allows us to propose two immunopathological stages of pulmonary COVID-19.

The segregation of autopsy lung samples from COVID-19 patients in two groups based on ISG expression contributes to our understanding of the interferon response against SARS-CoV-2. Like other coronaviruses, SARS-CoV-2 is particularly sensitive to type I interferons^8,26^. Therefore, and similar to other coronaviruses^27^, it has evolved strategies to evade the interferon response, and SARS-CoV-2 leads to relatively weak IFN-I/III release in host cells at low multiplicities of infection^8^. This initial delay of IFN-I/III production may facilitate initial virus replication in lungs, as studies with SARS-CoV in mice have shown, and enable an eventual increase of the IFN-I response and death^15^. A similar observation was made for fatal SARS-CoV infections in humans, which were accompanied by elevated expression of ISGs^28^. Since the SARS-CoV-2 receptor ACE2 is itself an ISG on lung epithelial cells^29^, virus infection and the interferon response may promote each other in this phase of the infection. This may explain the observed correlation of high ISG expression and high viral load in lungs and widespread presence of SARS-CoV-2 in lung epithelial cells. Together this may contribute to fatal outcome of SARS-CoV-2 infections in the ISG^high^ group.

The observation of the ISG^high^ pattern in COVID-19 autopsy lungs seems to be at odds with initial observations that critical COVID-19 patients express on average lower ISGs in blood than patients with a milder course of disease^30^. One possible explanation is that the blood ISG status is not directly reflective of the lung. In support of this idea, bronchoalveolar lavages (BALs) from severe/critical patients showed high proportions of ISG^high^ macrophages and high expression of CXCL9/10/11, IL6, IL-1b, TNF, CCL2^4^, which is reflective of our ISG^high^ phenotype in lung autopsies. An alternative explanation for the apparent disconnect of lung and blood ISG status may come from the overall frequency of the ISG^high^ subtype of critical/fatal COVID-19: ISGs are highly expressed in blood of COVID-19 patients during the early, innate phase of COVID-19^9^, and we show that patients with an ISG^high^ status in lungs die early upon hospitalization. While the early-mortality subset accounted for 44% of all deaths in our study, epidemiological data from France^24^ suggest that this early critical subset is actually smaller: only 15% of patients died early after hospitalization. This percentage is consistent with the study by Hadjadj *et al*., which detects high ISG expression in blood of 3/17 (18%) critical COVID-19 patients, yet this signal gets diluted in the majority of ISG^low^ critical cases^30^. These two alternative explanations show how critical it will be to compare gene expression in blood and lungs of individual patients at different times of the infection and to identify peripheral biomarkers for COVID-19 lung status.

ISG^low^ COVID-19 patients in our study die with classical features of DAD^31^, on average 9.1 days after hospitalization. Later death compared to patients with an ISG^high^ pattern and progressive decline of systemic ISG expression during COVID-19^9,13^ led us to infer that the ISG^low^ pattern in lungs reflects a later phase of pulmonary COVID-19. ISG^low^ lungs show higher frequencies of T and B lymphocytes, compared to ISG^high^ lungs. None of our fatal cases showed lung lymphocyte counts below control levels. Therefore COVID-19 associated lymphopenia in blood^23,32^ or spleens^3^ does not translate into lymphocyte depletion in infected lungs. Potential reasons are that the infected lung acts as a potent sink for circulating lymphocytes and that local proliferation limited recruitment from the blood, as was shown for CD8+ T cells in BAL of severe patients^4^. Consistent with previous observations^33^ we describe an activated CD8+ T cell signature in lungs of ISG^low^ patients that contain low viral counts. This suggests that CD8+ T cells are critical for antiviral protection, and may transition into a protective memory pool, as observed for SARS-CoV^34,35^. In addition, we found elevated frequencies of CD8+PD1+ cells in ISG^low^ lungs compared to ISG^high^ lungs, but not above control levels. The observation that PD-1 levels are elevated in peripheral CD8+ T cells of severe COVID-19 infection, and whether this indicates exhaustion, remains controversial^13,23^. Overall, although we did not have paired serum antibody levels available, the infiltration pattern of ISG^low^ lungs suggested adaptive immune activation.

While our study sheds further light on COVID-19 lung disease, conclusions on therapy must be drawn with caution. We found that early after hospitalization, ISG^high^ autopsy lungs had uniformly high titers of SARS-CoV-2, and others found that viral loads in swabs and sputum are highest in early COVID-19^36^. This could indicate that treatment with compounds that directly interfere with the SARS-CoV-2 replication cycle, e.g. protease or polymerase inhibitors, should start early. However, high expression of ISGs in some lung autopsies raises caution about the use of IFN-I/III as therapeutics, at least as long as causes and consequences of interferon signaling in COVID-19 lungs remain unclear. We found reduced viral counts in ISG^low^ patients, but did not identify the moment at which the body is cleared of the virus. Therefore our findings of reduced viral loads in ISG^low^ patients should not be taken as justification to withhold compounds that directly interfere with the SARS-CoV-2 replication cycle from patients. Extending previous work^37^, we found signs of elevated complement activation specifically in ISG^low^ lungs. However, it is not known whether complement is synchronously activated in patient lungs and plasma^38^. Therefore, our results do not provide a further step towards personalized patient care but strengthen the hypothesis that complement inhibitors may show therapeutic benefit, at least in some COVID-19 patients.

Our study has several limitations. The fact that patients with the innate / ISG^high^ stage die early while patients with the lymphocytic / ISG^low^ profile die late after hospitalization, together with knowledge about the immune reaction against other coronaviruses, strongly suggests that COVID-19 lung disease progresses from an ISG^high^ to an ISG^low^ stage. However, an autopsy study is, by design, not longitudinal. Therefore we do not have formal proof that all COVID-19 infected ISG^low^ lungs have undergone a previous ISG^high^ stage. Also, it is unknown why some patients die early and others late. In addition, our focus on the lung only allowed us to investigate pulmonary factors of patient mortality, i.e. an overshooting innate immune activation with IAH in both ISG^high^ and ISG^low^ cases and DAD in ISG^low^ cases. No reported cause of death was enriched in ISG^high^ or ISG^low^ patients, and multi-organ failure was reported as a cause of death only for two of our patients. Another limitation is that we lack gene expression data from blood of autopsy patients or serological data at the time of death. Therefore we were not able to identify peripheral biomarkers predicting specific immunological profiles in the lung. Finally, we analyzed our lungs with a focused gene expression set since quality and quantity of autopsy-derived RNA is often insufficient for unbiased methods. In spite of this technical limitation, which restricted our analysis, we were able to uncover two novel and distinct immunopathological profiles in lungs of fatal COVID-19.

Taken together, our autopsy study sheds light on two distinct courses of lethal COVID-19 in lungs. It remains to be seen whether interferon signaling is only associated with or causally involved in these disease courses. However, our study strengthens the notion that interferon signaling is a central determinant of the pulmonary immune response against SARS-CoV-2.

## Methods

### Ethics statement

This study was conducted according to the principles expressed in the Declaration of Helsinki. Ethics approval was obtained from the Ethics Committee of Northwestern and Central Switzerland (Project-ID 2020-00629). For all patients, either personal and/or family consent was obtained for autopsy and sample collection.

### Patients and sample collection

The study is based on the analysis of 16 out of 21 consecutive COVID-19 autopsies performed between March 9^th^ and April 14^th^ 2020 at the Institute of Pathology Liestal and Institute of Medical Genetics and Pathology Basel, Switzerland. Clinical features including symptoms, course of disease, comorbidities, laboratory results and therapy are listed in **Table 1a**. Detailed autopsy findings for each patient were recently published, and the identifiers (with the prefix “C”) for each COVID-19 patient are consistent with the description of this Swiss COVID-19 autopsy cohort^6^. In this study, we analysed formalin fixed and paraffin embedded (FFPE) lung tissue of distinct areas of the lungs of 16 of these COVID-19 patients. All 16 COVID-19 patients had positive nasopharyngeal swabs collected while alive. In all COVID-19 patients, diagnosis was confirmed by detection of SARS-CoV-2 in postmortal lung tissues. 5/16 patients were additionally tested by postmortal nasopharyngeal swabs which were positive for SARS-CoV-2 in all 5 cases.

As a control cohort, we selected 6 autopsies performed between January 2019 and March 2020 at the Institute of Pathology Liestal (“normal” patients N1 – N6). These control patients died of other, non-infectious causes and had a similar age, gender and cardiovascular risk profile. Patients with infections were excluded from this control cohort. Another control cohort consisted of 4 autopsies of patients suffering from various infections mainly with bacteria affecting the lung (patients with lung pathology, P1 – P4). Details for both control cohorts are listed in **Table 1b,c**. SARS-CoV-2 was ruled out for each control patient by PCR-examination of lung tissue samples.

### Nucleic acid extraction

RNA was extracted from up to six sections of FFPE tissue blocks using RecoverAll Total Nucleic Acid Isolation Kit (Cat No. AM1975, Thermo Fisher Scientific, Waltham, MA, USA). Extraction of DNA from up to 10 sections of FFPE tissue samples was automated by EZ1 Advanced XL (Qiagen, Hilden, Germany) using the EZ1 DNA Tissue Kit (Cat No. 953034, Qiagen, Hilden, Germany). Concentration of DNA and RNA were measured with Qubit 2.0 Fluorometer and Qubit dsDNA HS Assay or Qubit RNA HS Assay (Cat No. Q33230 & Q32852, Thermo Fisher Scientific, Waltham, MA, USA), respectively.

### Quantification of SARS-CoV-2 in FFPE tissue samples

*Post mortem* viral load was individually measured in all lung tissue blocks from all patients included in this study. SARS-CoV-2 was detected in 15ng of human total RNA using the TaqMan 2019-nCoV Assay Kit v1 (Cat No. A47532, Thermo Fisher Scientific, Waltham, MA, USA), which targets three genomic regions (ORFab1, S Protein, N Protein) specific for SARS-CoV-2 and the human RNase P gene (RPPH1). The copy numbers of the SARS-CoV-2 viral genome was determined by utilizing the TaqMan 2019-nCoV Control Kit v1 (Cat No. A47533, Thermo Fisher Scientific, Waltham, MA, USA) and a comparative “ΔΔC?” method. The control kit contains a synthetic sample with a defined amount of target molecules for the human RPPH1 and the three SARS-CoV-2 assays, and was re-analyzed in parallel with patient samples. For each patient sample, this method resulted in individual copy numbers of the human RPPH1 and the three SARS-CoV-2 targets. Finally, the mean copy number of the SARS-CoV-2 targets was normalized to 1 × 10^6^ RPPH1 transcripts.

### Profiling of immune response by targeted RNAseq

The expression levels of 398 genes, including genes relevant in innate and adaptive immune response and housekeeping genes for normalization, were analyzed with the Oncomine Immune Response Research Assay (OIRRA, Cat No. A32881, Thermo Fisher Scientific, Waltham, MA, USA). The OIRRA is a targeted gene expression assay designed for the Ion™ next-generation sequencing (NGS) platform. Our study focused on the analysis of rare autopsy tissue samples from COVID-19 patients collected in clinical routine during the COVID-19 pandemic. An inherent problem for transcriptomic studies of autopsy tissues is that it is often not possible to extract high quality RNA in sufficient amounts. To avoid sample dropout due to these reasons, we decided to use a robust and straightforward targeted gene expression assay (OIRRA) rather than whole transcriptome analysis. Since the focus of our study was to investigate the immune profile of lungs, an immunoprofiling assay was deemed most appropriate. The OIRRA gene expression assay was originally designed to interrogate the tumor microenvironment to enable mechanistic studies and identification of predictive biomarkers for immunotherapy in cancer. The assay is optimized to measure the expression of genes involved in immune cell interactions and signaling, including genes expressed at low levels and involved in inflammatory signaling. The 398 genes covered by this assay are listed in **Supplementary Table 2**. The accessibility of such commercially available assays could be an encouragement to hospitals around the world to conduct similar molecular profiling studies of diagnostic tissue samples from COVID-19 patients, allowing relatively fast and easy stratification of patients into distinct biological groups as a starting point for targeted intervention strategies.

The NGS libraries were prepared as recommended by the supplier. In brief, 30ng of total RNA were used for reverse transcription (SuperScript VILO, Cat No. 11754250, Thermo Fisher Scientific, Waltham, MA, USA) and subsequent library preparation. The libraries were quantified (Ion Library TaqMan Quantitation Kit, Cat No. 4468802, Thermo Fisher Scientific, Waltham, MA, USA), equimolarly pooled and sequenced utilizing the Ion GeneStudio S5xl (Thermo Fisher Scientific, Waltham, MA, USA). De-multiplexing and gene expression level quantification were performed with the standard setting of the ImmuneResponseRNA plugin (version 5.12.0.1) within the Torrent Suite (version 5.12.1), provided as part of the OIRRA by Thermo Fisher Scientific, Waltham, MA, USA.

### Detection of co-infections by whole genome sequencing

To identify potential pathogens accompanying an infection with SARS-CoV-2, we analyzed the DNA of the same tissue samples used for detection and profiling of the SARS-CoV-2-specific immune response. First, 250ng of genomic DNA was enzymatically sheared (15 minutes at 37°C) and barcoded using the Ion Xpress Plus Fragment Library Kit (Cat No. 4471269, Thermo Fisher Scientific, Waltham, MA, USA). Subsequently, the libraries were quantified (Ion Library TaqMan Quantitation Kit, Cat No. 4468802, Thermo Fisher Scientific, Waltham, MA, USA) and up to three libraries were pooled at equimolar levels for analysis with Ion GeneStudio S5xl (Thermo Fisher Scientific, Waltham, MA, USA). Sequencing data for each sample was analysed using the CLC genomics workbench (version 20.0.3, Qiagen, Hilden, Germany) in combination with the microbial genomics module (version 20.0.1, Qiagen, Hilden, Germany): The raw reads were trimmed by quality (Mott algorithm with limit 0.05 and a maximum of 2 ambiguous bases per read) and mapped to the human genome (GRCh37 hg19, match score: 1, mismatch cost: 2, indel opening cost: 6, indel extension cost: 1). Unmapped reads were analysed by taxonomic profiling to identify reads of viral or bacterial origin. The profiling utilized an index of 11’540 viral genomes with a minimum length of 1’000 bp and 2’715 bacterial reference genomes with a minimum length of 500’000 bp, retrieved from the NCBI Reference Sequence Database (date of download: 2020-04-02).

### Immunohistochemistry

Immunohistochemical analyses for CD3, CD4, CD8, CD15, CD20, CD68, CD123, CD163, PD-1, MPO, p53, Ki67, C3d and C5b-9 were performed on all lung tissue blocks used in this study. Antibodies, staining protocols and conditions are detailed in **Supplementary Table 4**.

### Qualitative and semiquantitative assessment of histopathological lung damage and neutrophilic infiltration

Hematoxylin and eosin (H&E) and Elastica van Gieson (EvG) stained sections of all lung tissues used in this study were independently evaluated by two experienced and board certified pathologists (VZ and KDM) (**Supplementary Table 5**). Both pathologists evaluated the presence of diffuse alveolar damage (DAD), and if present, its stage, intra-alveolar edema and hemorrhage. The characteristic three phases or stages of DAD - exudative (1), proliferative / organizing (2), fibrotic (3) - were assessed as described^39^. In our cohort of COVID-19 lungs, we observed only DAD stages 1 and/or 2, and the fibrotic phase (3) was not observed. In addition, both pathologists evaluated the severity of histopathological changes in COVID-19 lungs (1 = mild / discrete alterations, 2 = moderate, 3 = severe changes) based on resemblance between normal and pathologically altered lung tissues. Parameters that were taken into account included reduction of alveolar air-filled spaces, typical histologic features of DAD with hyaline membrane formation, infiltration of lymphocytes, monocytes and neutrophils into interstitial and alveolar spaces, type 2 pneumocyte hyperplasia, desquamation of pneumocytes, histologic features of organizing pneumonia including intra-alveolar fibrin deposition and fibrosis (acute fibrinous and organizing pneumonia, AFOP)^40,41^. The number of neutrophils per lung tissue section was estimated on H&E stained sections and by immunohistochemical stains for CD15 and MPO using a three tiered system (1 = few or no neutrophils, 2 = moderate number of neutrophils, 3 = high number of neutrophils). Assessment of the two pathologists was concordant in the vast majority of cases. Discrepant cases were reviewed by a third pathologist (NW) to reach consent.

### Digital image analysis

Slides were digitalized on a 3DHistech™ P1000 slide scanner at 400x magnification (3DHISTECH Ltd. Budapest, Hungary). Digital slide review and quality control was performed by a board-certified pathologist (VHK). Tissue regions with staining artefacts, folds or other technical artefacts were excluded from analysis. A deep neural network (DNN) algorithm (Simoyan and Zisserman VGG, HALO AI™ on HALO™ 3.0.311.167, Indica Labs, Corrales, NM) was trained using pathologist annotations to automatically localize and measure the area of each lung tissue sample on the digital slides. Background regions and glass were excluded from analysis. Mark-up images for tissue classification were generated and classification accuracy was confirmed through pathology review. For cell-level analysis, color deconvolution for DAB, AP and hematoxylin channels was performed and nuclear segmentation was optimized using cell-morphometric parameters. Marker-positive cells in stromal and epithelial regions were quantified. For CD3, CD4, CD8, CD20, CD68, CD123, CD163 and PD1, staining detection was optimized for the cytoplasmic / membranous compartment and marker expression was measured on a continuous scale at single cell resolution. For assessment of CD8/PD1 double stains, color deconvolution was optimized for separation of DAB (PD1) and AP (CD8) staining products. Internal controls (non-immune cells) and external controls (tonsil) were used to calibrate the detection limits and cross-validated by visual review. For each tissue sample, the total area of lung tissue in mm^2^, the absolute number of marker-positive cells, cell morphometric parameters and staining intensity were recorded.

### Identification of SARS-CoV-2 immune response pattern

#### Gene expression analysis

Samples were included in the study based on quality of libraries and alignment performance. Applied inclusion criteria are: >1 million of mapped reads, good concentration of libraries, average read length >100bp, > 300 target genes with more than 10 reads. One sample with > 1 Mio reads was excluded from the study because of shorter read length and a low library concentration. Notably this sample had the longest time between death and autopsy (72h) before analysis. Differential expression analysis was performed using the edgeR package comparing normal lung samples, COVID-19 samples and samples from patients with other infections. Genes were selected for downstream analyses by fdr <0.05 and |logFC| >1 for clustering analysis. Clustering analysis was performed using k-means algorithm and complete linkage. Ideal number of clusters (n=3) was chosen based on 30 different algorithms^42^ and the final clustering derives from the consensus of 2000 iterations. Expression of gene signatures was calculated as median of log2(cpm + 1) of selected genes.

#### Functional enrichment analysis

Biological processes enrichment was performed using the enrichGO function of the package clusterProfiler^43^ setting all the genes included in the assay as universe.

#### Statistical analysis

All the analyses and graphical representations were performed using the R statistical environment software^44^ and the following packages: ggplot2^45^, circlize^46^, ComplexHeatmap^47^, ggfortify^48^, reshape2^49^ and factoextra^50^. Correlation between transcripts and viral counts was performed using Pearson’s correlation. Association between continuous and categorical data were tested using Wilcoxon rank sum test.

Based on the often non-uniform histopathological appearance of lung samples from the same patient, transcriptomic, morphologic or histopathological analyses were performed at the tissue sample level. Analyses involving patients’ clinical or demographical data were performed at the patient level, and patients in which all analyzed lung samples expressed an ISG^high^ or an ISG^low^ profile were called ISG^high^ or ISG^low^ patients.

Box-plots elements indicate the median (center line), upper and lower quartiles (box limits) and show all the data points. Whiskers extend to the most extreme value included in 1.5x interquartile range.

## Data Availability

The datasets generated and analysed during this study can be accessed in GEO (GEO Submission (GSE151764) [NCBI tracking system #20999475]) and are available from the corresponding author upon request.

## Data availability

The datasets generated and analysed within this study can be accessed in GEO (GSE151764) and are available from the corresponding author upon request.

## Acknowledgements

VHK gratefully acknowledges funding by the Promedica Foundation (F-87701-41-01). AT, JDH, TM and KDM are supported by and gratefully acknowledge the Botnar Research Centre for Child Health. We would like to thank Christian Tosch, Beat Béni, Daniel Turek, Melanie Sachs, Anne Graber, Christina Herz, Arbeneshe Berisha, Norbert Wey and the USZ pathology IT team, André Fitsche, Marcel Glönkler, Christiane Mittman and the USZ pathology laboratory team for expert technical support and scanning of slides.

## Author contributions

RN, YC, VHK, FD, TJ and KDM jointly conceived the study, performed data interpretation and prepared the manuscript. AT, MB, HM, MT and CA provided intellectual input, provided critical resources and critically reviewed the manuscript. RN, YC, VHK, MH, TH and TJ performed bioinformatic and statistical analysis. AT, JDH, TM, NS, AF collected autopsy specimens, patient data and performed experiments. VZ, NW, WK and CA performed histomorphological evaluation. All authors approved the final manuscript.

## Competing interests

VHK has served as an invited speaker on behalf of Indica Labs. TH and TJ are employees of Novartis. The other authors declare no competing interests.

**Supplementary Table 2.**
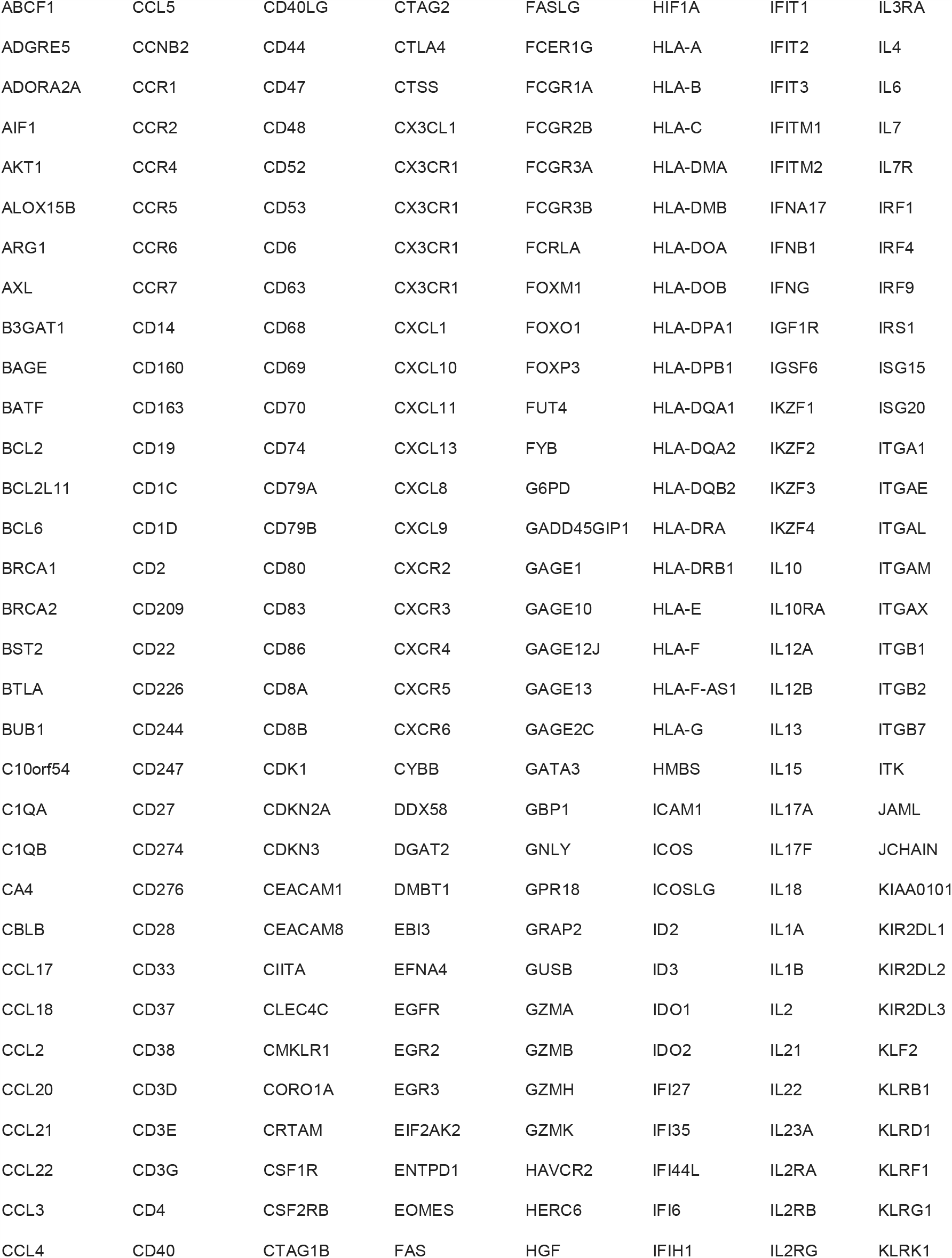
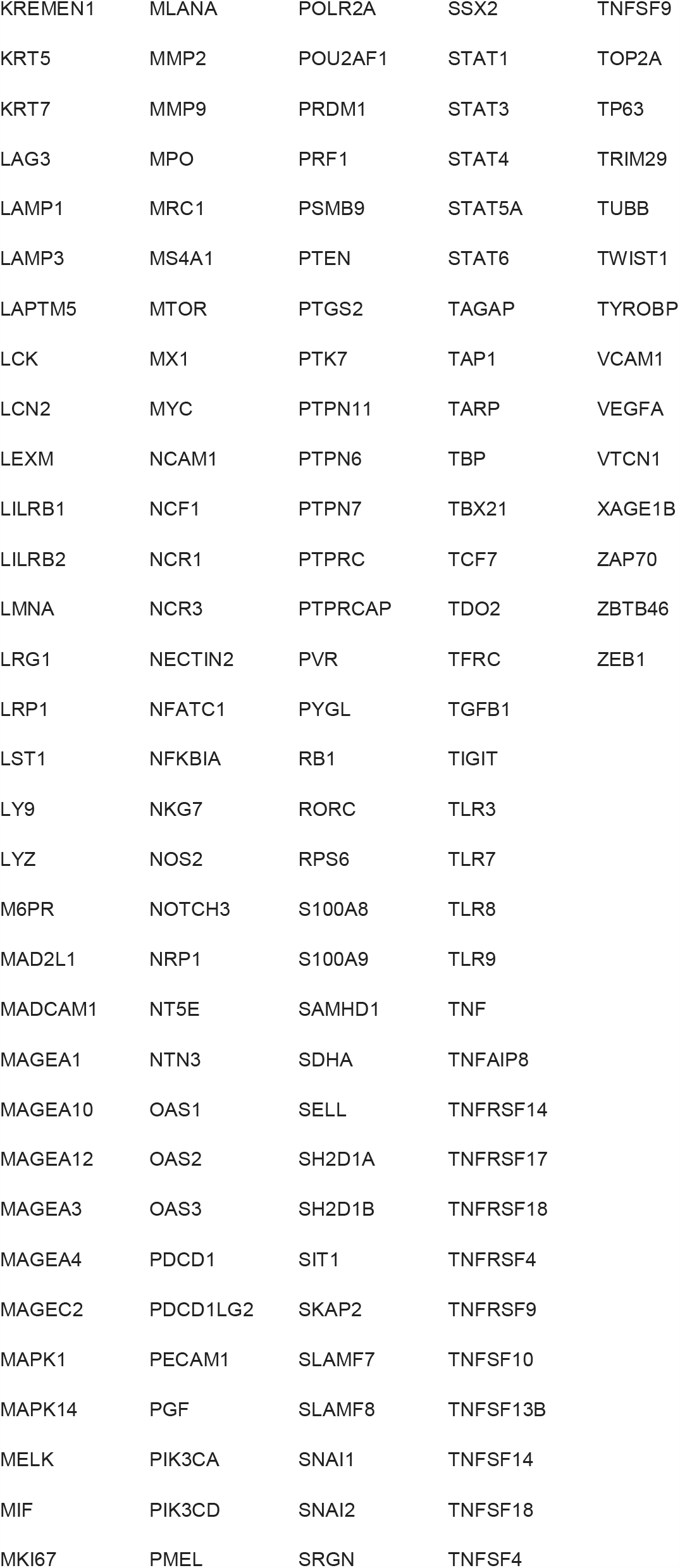
OIRRA gene list.

|  |  |  |  |  |  |  |  |
| --- | --- | --- | --- | --- | --- | --- | --- |
| ABCF1 | CCL5 | CD40LG | CTAG2 | FASLG | HIF1A | IFIT1 | IL3RA |
| ADGRE5 | CCNB2 | CD44 | CTLA4 | FCER1G | HLA-A | IFIT2 | IL4 |
| ADORA2A | CCR1 | CD47 | CTSS | FCGR1A | HLA-B | IFIT3 | IL6 |
| AIF1 | CCR2 | CD48 | CX3CL1 | FCGR2B | HLA-C | IFITM1 | IL7 |
| AKT1 | CCR4 | CD52 | CX3CR1 | FCGR3A | HLA-DMA | IFITM2 | IL7R |
| ALOX15B | CCR5 | CD53 | CX3CR1 | FCGR3B | HLA-DMB | IFNA17 | IRF1 |
| ARG1 | CCR6 | CD6 | CX3CR1 | FCRLA | HLA-DOA | IFNB1 | IRF4 |
| AXL | CCR7 | CD63 | CX3CR1 | FOXM1 | HLA-DOB | IFNG | IRF9 |
| B3GAT1 | CD14 | CD68 | CXCL1 | FOXO1 | HLA-DPA1 | IGF1R | IRS1 |
| BAGE | CD160 | CD69 | CXCL10 | FOXP3 | HLA-DPB1 | IGSF6 | ISG15 |
| BATF | CD163 | CD70 | CXCL11 | FUT4 | HLA-DQA1 | IKZF1 | ISG20 |
| BCL2 | CD19 | CD74 | CXCL13 | FYB | HLA-DQA2 | IKZF2 | ITGA1 |
| BCL2L11 | CD1C | CD79A | CXCL8 | G6PD | HLA-DQB2 | IKZF3 | ITGAE |
| BCL6 | CD1D | CD79B | CXCL9 | GADD45GIP1 | HLA-DRA | IKZF4 | ITGAL |
| BRCA1 | CD2 | CD80 | CXCR2 | GAGE1 | HLA-DRB1 | IL10 | ITGAM |
| BRCA2 | CD209 | CD83 | CXCR3 | GAGE10 | HLA-E | IL10RA | ITGAX |
| BST2 | CD22 | CD86 | CXCR4 | GAGE12J | HLA-F | IL12A | ITGB1 |
| BTLA | CD226 | CD8A | CXCR5 | GAGE13 | HLA-F-AS1 | IL12B | ITGB2 |
| BUB1 | CD244 | CD8B | CXCR6 | GAGE2C | HLA-G | IL13 | ITGB7 |
| C10orf54 | CD247 | CDK1 | CYBB | GATA3 | HMBS | IL15 | ITK |
| C1QA | CD27 | CDKN2A | DDX58 | GBP1 | ICAM1 | IL17A | JAML |
| C1QB | CD274 | CDKN3 | DGAT2 | GNLY | ICOS | IL17F | JCHAIN |
| CA4 | CD276 | CEACAM1 | DMBT1 | GPR18 | ICOSLG | IL18 | KIAA0101 |
| CBLB | CD28 | CEACAM8 | EBI3 | GRAP2 | ID2 | IL1A | KIR2DL1 |
| CCL17 | CD33 | CIITA | EFNA4 | GUSB | ID3 | IL1B | KIR2DL2 |
| CCL18 | CD37 | CLEC4C | EGFR | GZMA | IDO1 | IL2 | KIR2DL3 |
| CCL2 | CD38 | CMKLR1 | EGR2 | GZMB | IDO2 | IL21 | KLF2 |
| CCL20 | CD3D | CORO1A | EGR3 | GZMH | IFI27 | IL22 | KLRB1 |
| CCL21 | CD3E | CRTAM | EIF2AK2 | GZMK | IFI35 | IL23A | KLRD1 |
| CCL22 | CD3G | CSF1R | ENTPD1 | HAVCR2 | IFI44L | IL2RA | KLRF1 |
| CCL3 | CD4 | CSF2RB | EOMES | HERC6 | IFI6 | IL2RB | KLRG1 |
| CCL4 | CD40 | CTAG1B | FAS | HGF | IFIH1 | IL2RG | KLRK1 |

|  |  |  |  |  |
| --- | --- | --- | --- | --- |
| KREMEN1 | MLANA | POLR2A | SSX2 | TNFSF9 |
| KRT5 | MMP2 | POU2AF1 | STAT1 | TOP2A |
| KRT7 | MMP9 | PRDM1 | STAT3 | TP63 |
| LAG3 | MPO | PRF1 | STAT4 | TRIM29 |
| LAMP1 | MRC1 | PSMB9 | STAT5A | TUBB |
| LAMP3 | MS4A1 | PTEN | STAT6 | TWIST1 |
| LAPTM5 | MTOR | PTGS2 | TAGAP | TYROBP |
| LCK | MX1 | PTK7 | TAP1 | VCAM1 |
| LCN2 | MYC | PTPN11 | TARP | VEGFA |
| LEXM | NCAM1 | PTPN6 | TBP | VTCN1 |
| LILRB1 | NCF1 | PTPN7 | TBX21 | XAGE1B |
| LILRB2 | NCR1 | PTPRC | TCF7 | ZAP70 |
| LMNA | NCR3 | PTPRCAP | TDO2 | ZBTB46 |
| LRG1 | NECTIN2 | PVR | TFRC | ZEB1 |
| LRP1 | NFATC1 | PYGL | TGFB1 |  |
| LST1 | NFKBIA | RB1 | TIGIT |  |
| LY9 | NKG7 | RORC | TLR3 |  |
| LYZ | NOS2 | RPS6 | TLR7 |  |
| M6PR | NOTCH3 | S100A8 | TLR8 |  |
| MAD2L1 | NRP1 | S100A9 | TLR9 |  |
| MADCAM1 | NT5E | SAMHD1 | TNF |  |
| MAGEA1 | NTN3 | SDHA | TNFAIP8 |  |
| MAGEA10 | OAS1 | SELL | TNFRSF14 |  |
| MAGEA12 | OAS2 | SH2D1A | TNFRSF17 |  |
| MAGEA3 | OAS3 | SH2D1B | TNFRSF18 |  |
| MAGEA4 | PDCD1 | SIT1 | TNFRSF4 |  |
| MAGEC2 | PDCD1LG2 | SKAP2 | TNFRSF9 |  |
| MAPK1 | PECAM1 | SLAMF7 | TNFSF10 |  |
| MAPK14 | PGF | SLAMF8 | TNFSF13B |  |
| MELK | PIK3CA | SNAI1 | TNFSF14 |  |
| MIF | PIK3CD | SNAI2 | TNFSF18 |  |
| MKI67 | PMEL | SRGN | TNFSF4 |  |

**Supplementary Table 3.**
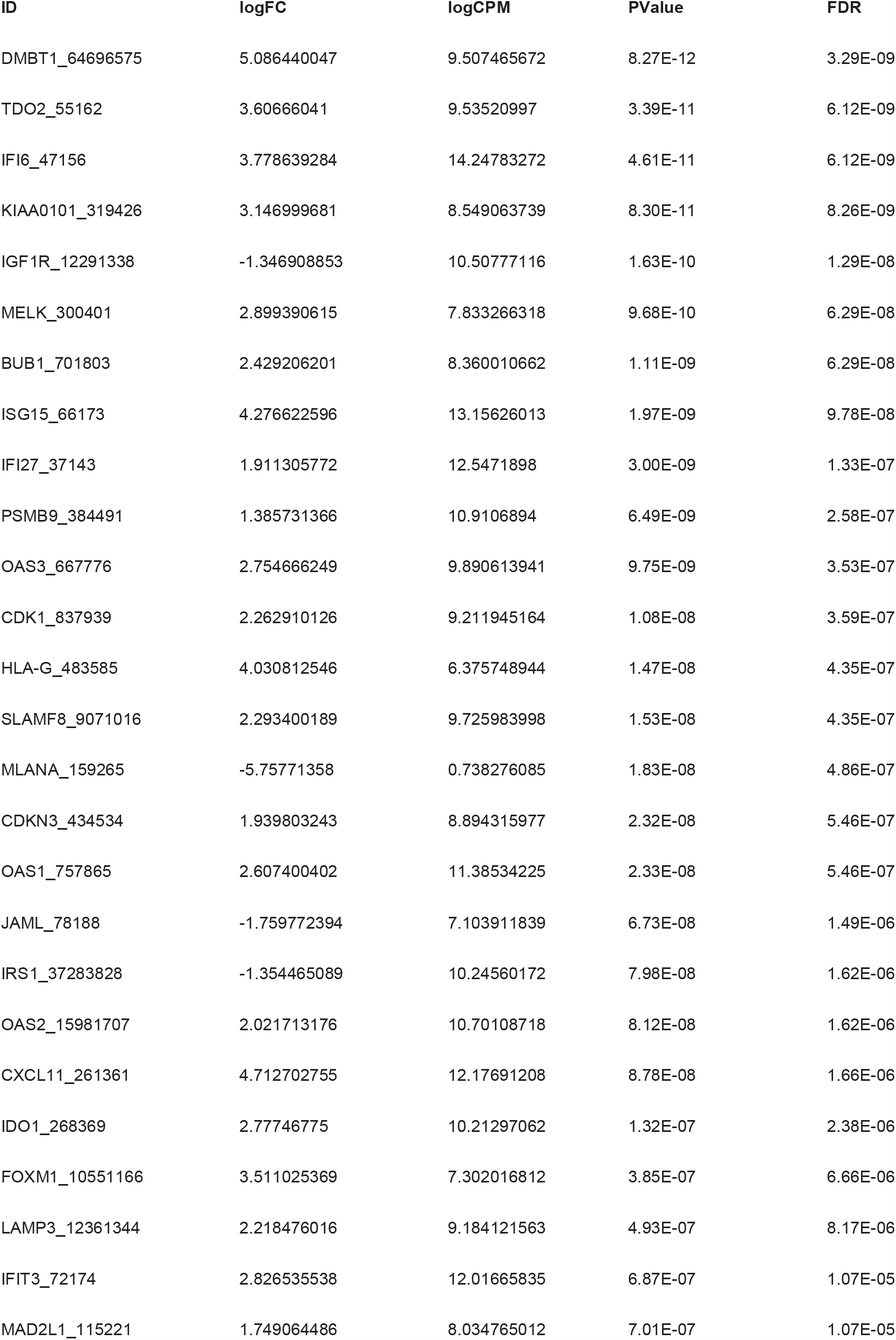
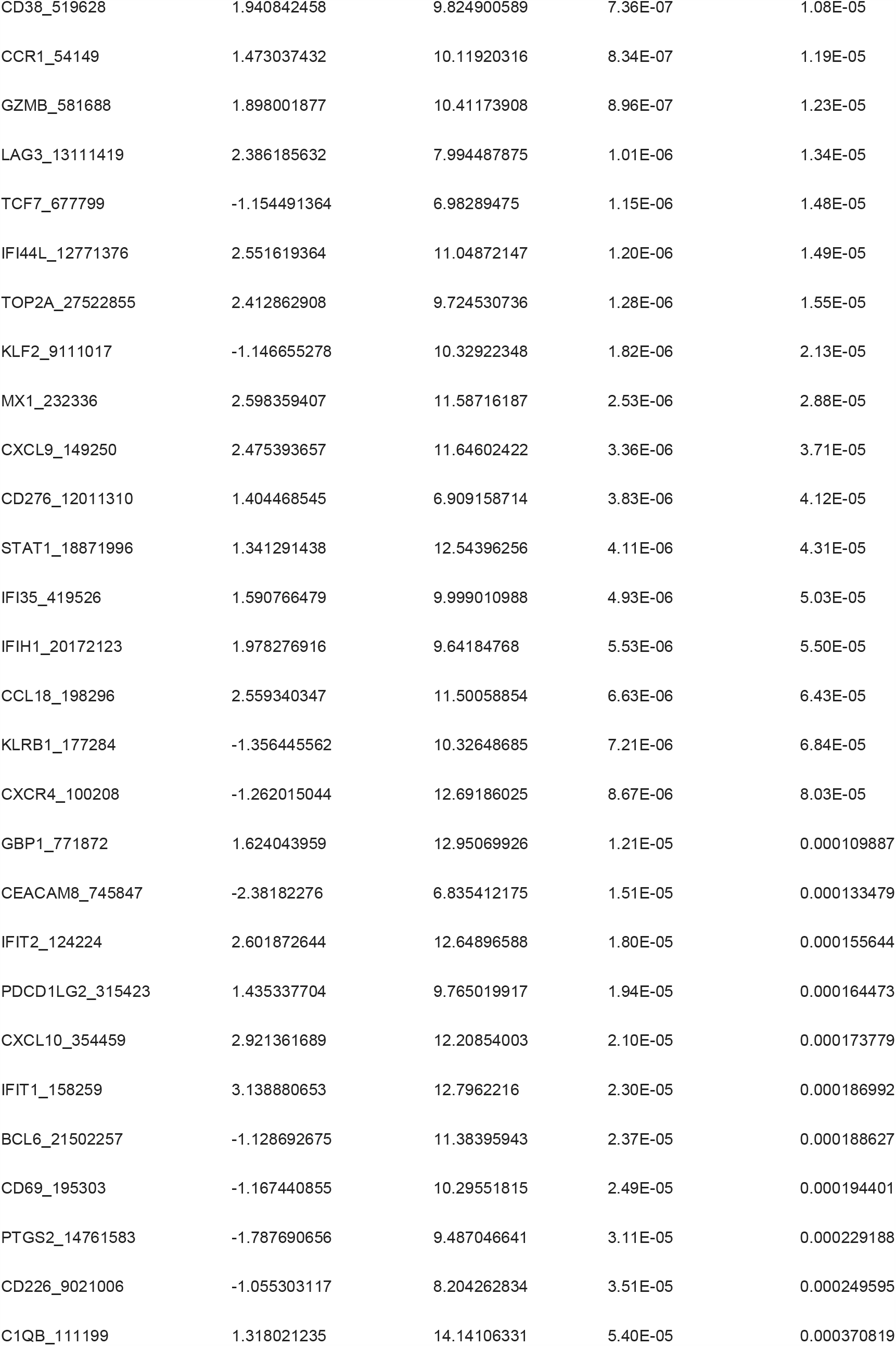
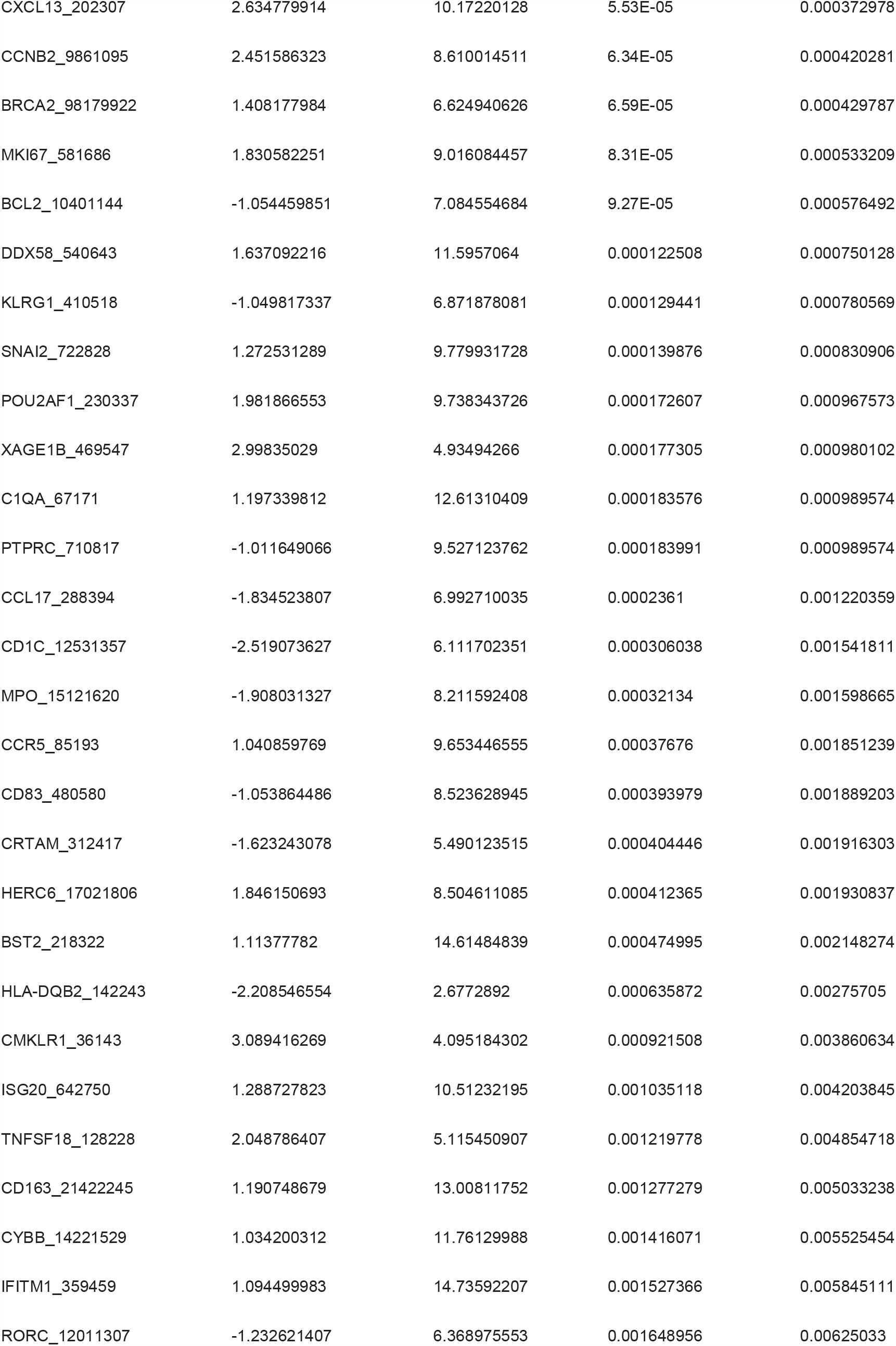
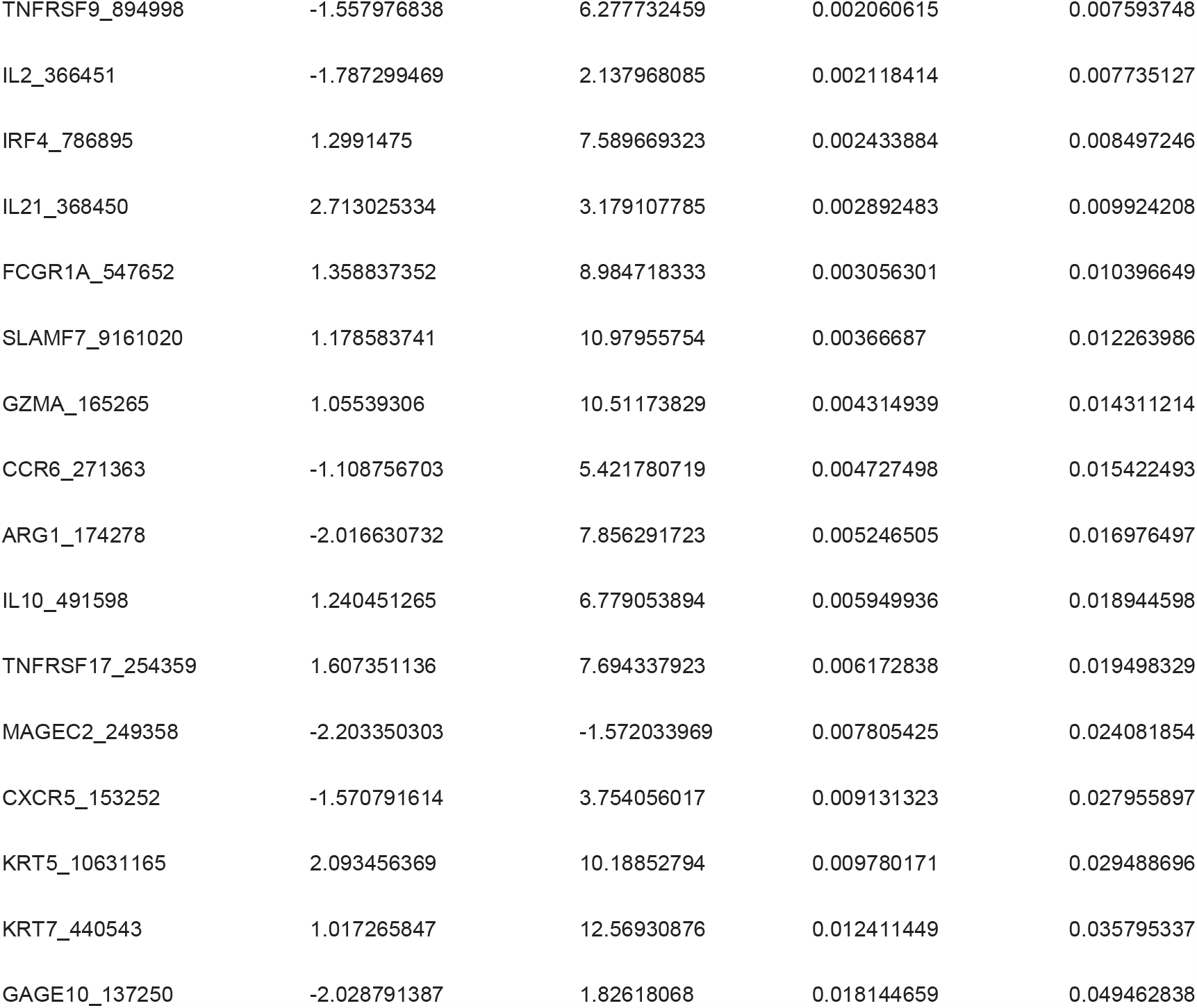
Differentially expressed genes, COVID-19 versus controls.

**Supplementary Table 4.**
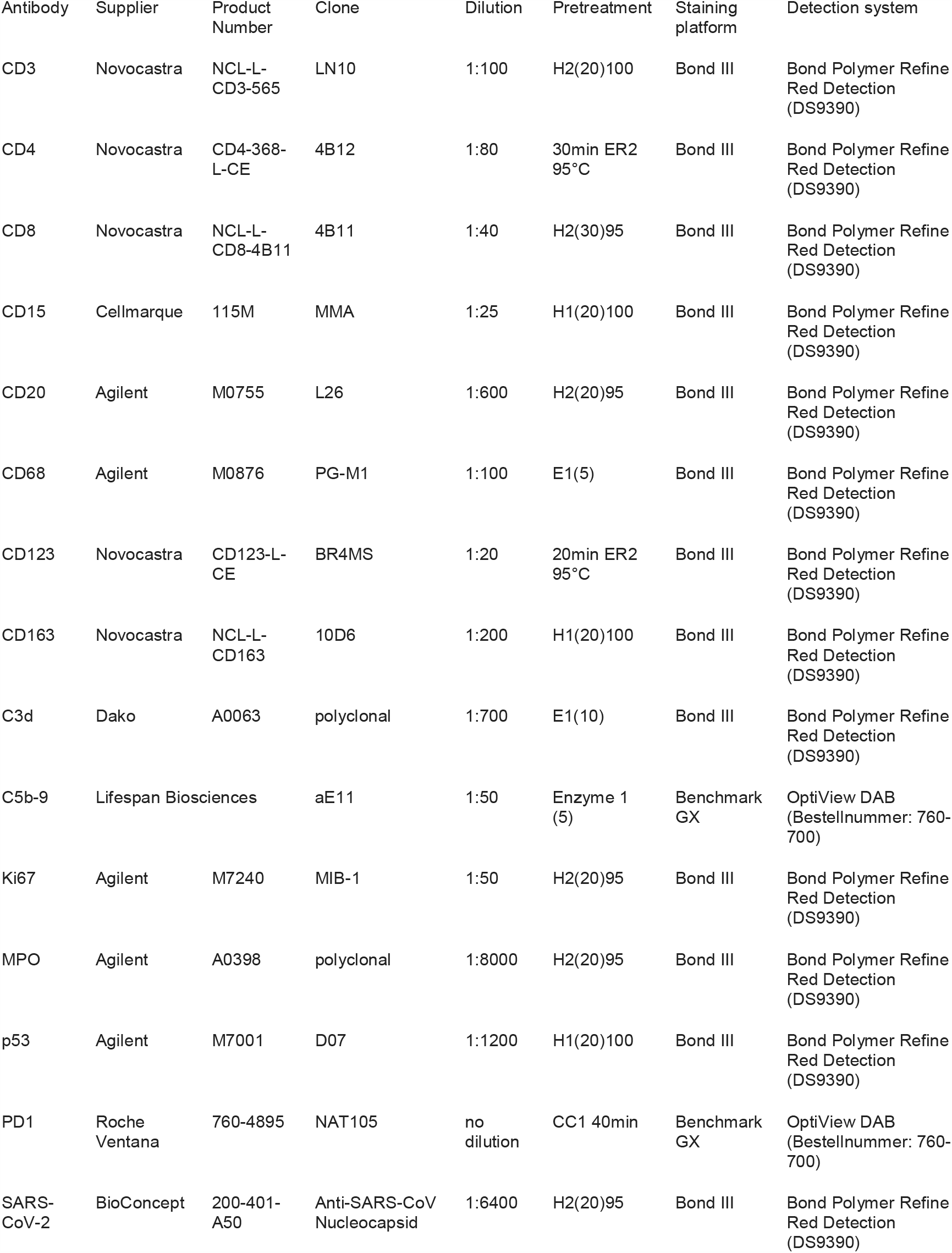
Antibodies and staining conditions.

**Supplementary Table 5.**
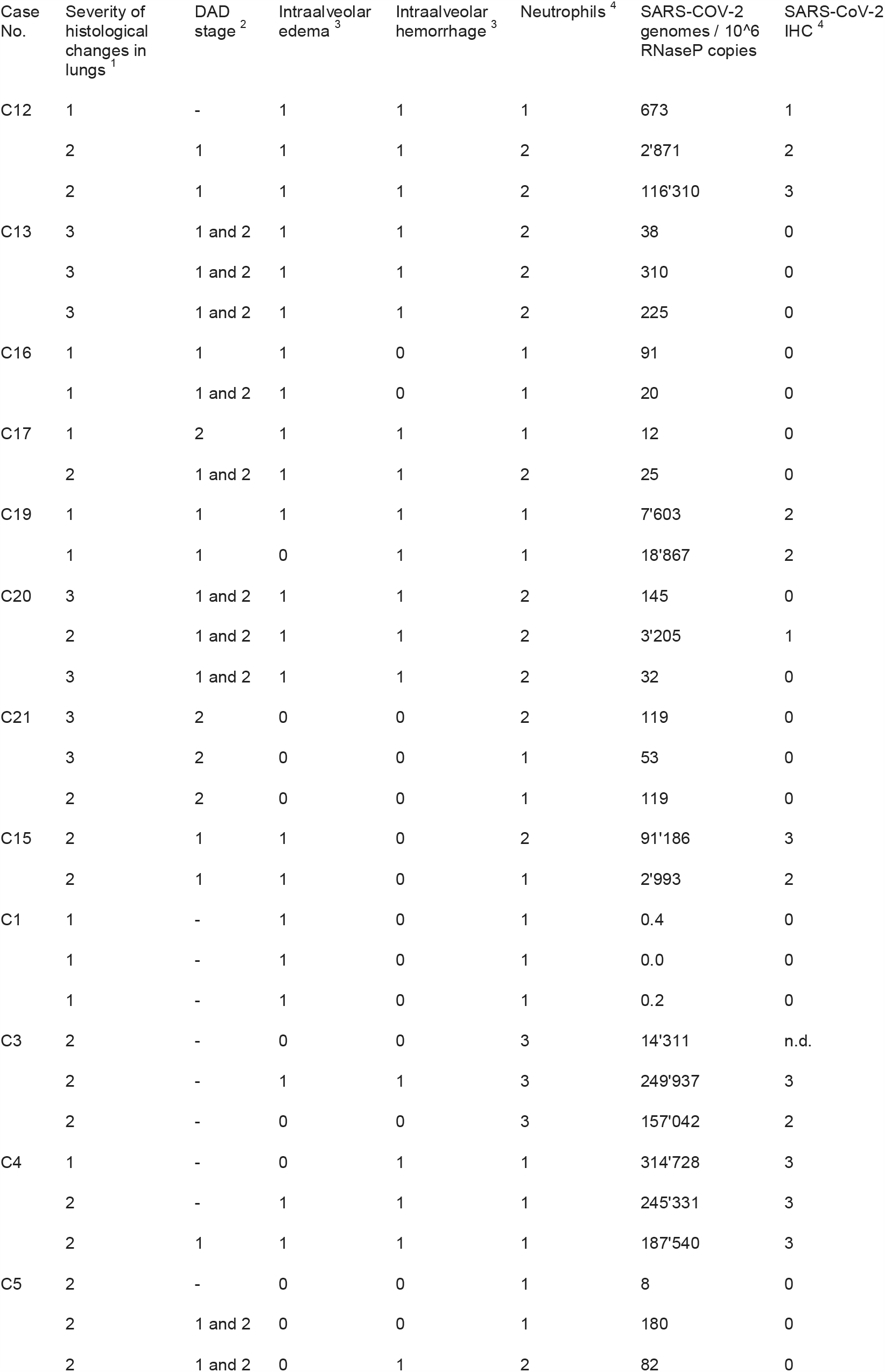
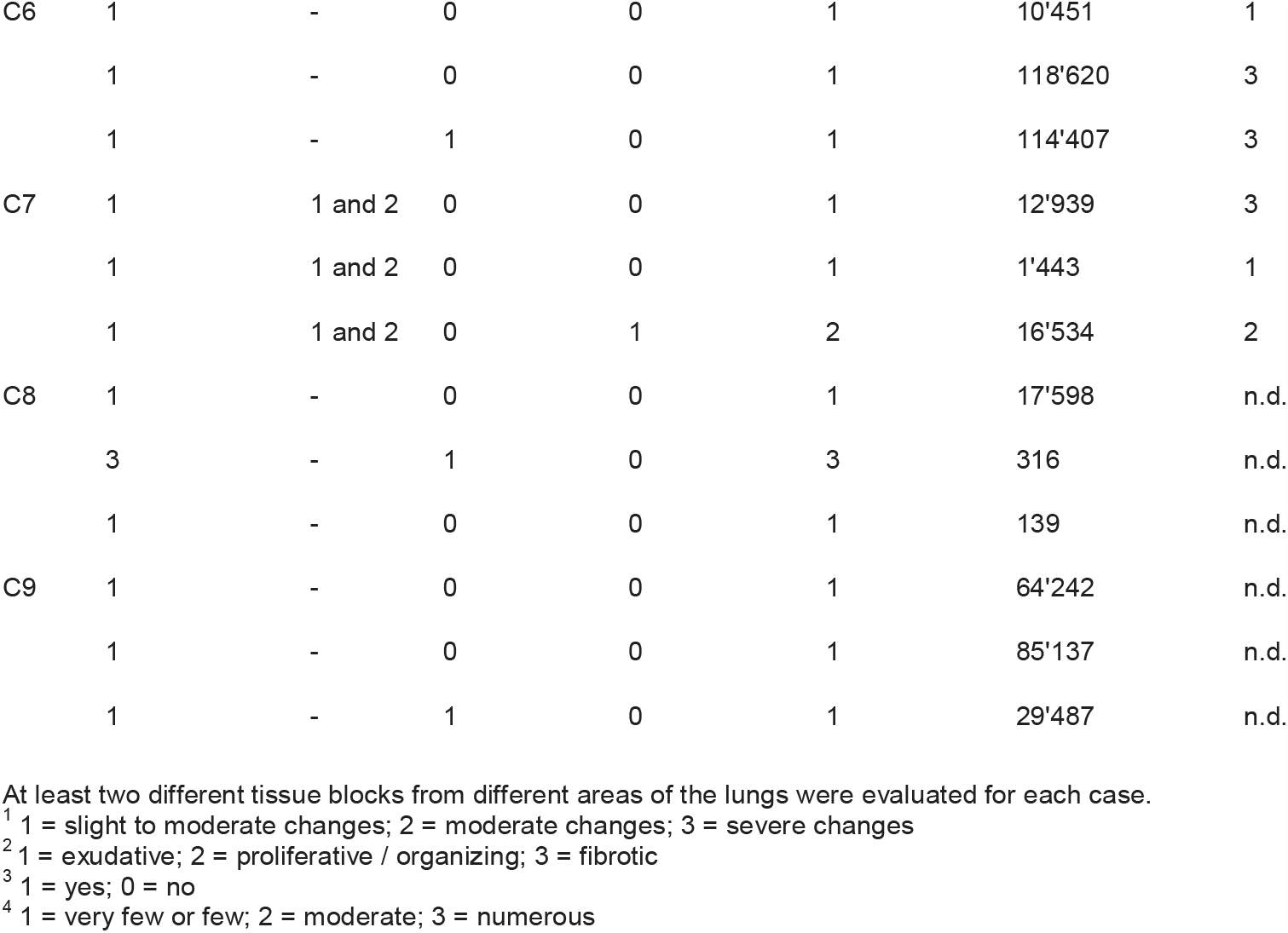
Histopathology.

| Case No. | Severity of histological changes in lungs<sup>1</sup> | DAD stage<sup>2</sup> | Intraalveolar edema<sup>3</sup> | Intraalveolar hemorrhage<sup>3</sup> | Neutrophils<sup>4</sup> | SARS-COV-2 genomes / 10<sup>6</sup> RNaseP copies | SARS-CoV-2 IHC<sup>4</sup> |
| --- | --- | --- | --- | --- | --- | --- | --- |
| C12 | 1 | - | 1 | 1 | 1 | 673 | 1 |
|  | 2 | 1 | 1 | 1 | 2 | 2'871 | 2 |
|  | 2 | 1 | 1 | 1 | 2 | 116'310 | 3 |
| C13 | 3 | 1 and 2 | 1 | 1 | 2 | 38 | 0 |
|  | 3 | 1 and 2 | 1 | 1 | 2 | 310 | 0 |
|  | 3 | 1 and 2 | 1 | 1 | 2 | 225 | 0 |
| C16 | 1 | 1 | 1 | 0 | 1 | 91 | 0 |
|  | 1 | 1 and 2 | 1 | 0 | 1 | 20 | 0 |
| C17 | 1 | 2 | 1 | 1 | 1 | 12 | 0 |
|  | 2 | 1 and 2 | 1 | 1 | 2 | 25 | 0 |
| C19 | 1 | 1 | 1 | 1 | 1 | 7'603 | 2 |
|  | 1 | 1 | 0 | 1 | 1 | 18'867 | 2 |
| C20 | 3 | 1 and 2 | 1 | 1 | 2 | 145 | 0 |
|  | 2 | 1 and 2 | 1 | 1 | 2 | 3'205 | 1 |
|  | 3 | 1 and 2 | 1 | 1 | 2 | 32 | 0 |
| C21 | 3 | 2 | 0 | 0 | 2 | 119 | 0 |
|  | 3 | 2 | 0 | 0 | 1 | 53 | 0 |
|  | 2 | 2 | 0 | 0 | 1 | 119 | 0 |
| C15 | 2 | 1 | 1 | 0 | 2 | 91'186 | 3 |
|  | 2 | 1 | 1 | 0 | 1 | 2'993 | 2 |
| C1 | 1 | - | 1 | 0 | 1 | 0.4 | 0 |
|  | 1 | - | 1 | 0 | 1 | 0.0 | 0 |
|  | 1 | - | 1 | 0 | 1 | 0.2 | 0 |
| C3 | 2 | - | 0 | 0 | 3 | 14'311 | n.d. |
|  | 2 | - | 1 | 1 | 3 | 249'937 | 3 |
|  | 2 | - | 0 | 0 | 3 | 157'042 | 2 |
| C4 | 1 | - | 0 | 1 | 1 | 314'728 | 3 |
|  | 2 | - | 1 | 1 | 1 | 245'331 | 3 |
|  | 2 | 1 | 1 | 1 | 1 | 187'540 | 3 |
| C5 | 2 | - | 0 | 0 | 1 | 8 | 0 |
|  | 2 | 1 and 2 | 0 | 0 | 1 | 180 | 0 |
|  | 2 | 1 and 2 | 0 | 1 | 2 | 82 | 0 |
| C6 | 1 | - | 0 | 0 | 1 | 10'451 | 1 |
|  | 1 | - | 0 | 0 | 1 | 118'620 | 3 |
|  | 1 | - | 1 | 0 | 1 | 114'407 | 3 |
| C7 | 1 | 1 and 2 | 0 | 0 | 1 | 12'939 | 3 |
|  | 1 | 1 and 2 | 0 | 0 | 1 | 1'443 | 1 |
|  | 1 | 1 and 2 | 0 | 1 | 2 | 16'534 | 2 |
| C8 | 1 | - | 0 | 0 | 1 | 17'598 | n.d. |
|  | 3 | - | 1 | 0 | 3 | 316 | n.d. |
|  | 1 | - | 0 | 0 | 1 | 139 | n.d. |
| C9 | 1 | - | 0 | 0 | 1 | 64'242 | n.d. |
|  | 1 | - | 0 | 0 | 1 | 85'137 | n.d. |
|  | 1 | - | 1 | 0 | 1 | 29'487 | n.d. |

At least two different tissue blocks from different areas of the lungs were evaluated for each case.
<sup>1</sup> 1 = slight to moderate changes; 2 = moderate changes; 3 = severe changes
<sup>2</sup> 1 = exudative; 2 = proliferative / organizing; 3 = fibrotic
<sup>3</sup> 1 = yes; 0 = no
<sup>4</sup> 1 = very few or few; 2 = moderate; 3 = numerous

## References

1. Hopkins, J. Corona virus resource center. Latest update: 05/22/2020): https://coronavirus.jhu.edu/data (2020).

2. Wu, Z. & McGoogan, J.M. Characteristics of and Important Lessons From the Coronavirus Disease 2019 (COVID-19) Outbreak in China: Summary of a Report of 72314 Cases From the Chinese Center for Disease Control and Prevention. JAMA (2020).

3. Chen, G., et al. Clinical and immunological features of severe and moderate coronavirus disease 2019. J Clin Invest 130, 2620–2629 (2020).

4. Liao, M., et al. Single-cell landscape of bronchoalveolar immune cells in patients with COVID-19. Nat Med 26, 842–844 (2020).

5. Vabret, N., et al. Immunology of COVID-19: Current State of the Science. Immunity 52, 910–941 (2020).

6. Menter, T., et al. Post-mortem examination of COVID19 patients reveals diffuse alveolar damage with severe capillary congestion and variegated findings of lungs and other organs suggesting vascular dysfunction. Histopathology (2020).

7. Charrad, M., Ghazzali, N., Boiteau, V. & Niknafs, A. NbClust Package: finding the relevant number of clusters in a dataset. J. Stat. Softw (2012).

8. Blanco-Melo, D., et al. Imbalanced Host Response to SARS-CoV-2 Drives Development of COVID-19. Cell 181, 1036–1045 e1039 (2020).

9. Yan, Q., et al. Longitudinal peripheral blood transcriptional analysis of COVID-19 patients captures disease progression and reveals potential biomarkers. medRxiv (2020).

10. Lieberman, N.A.P., et al. In vivo antiviral host response to SARS-CoV-2 by viral load, sex, and age. bioRxiv (2020).

11. Zhang, D., et al. COVID-19 infection induces readily detectable morphological and inflammation-related phenotypic changes in peripheral blood monocytes, the severity of which correlate with patient outcome. medRxiv (2020).

12. Huang, C., et al. Clinical features of patients infected with 2019 novel coronavirus in Wuhan, China. Lancet 395, 497–506 (2020).

13. Wilk, A.J., et al. A single-cell atlas of the peripheral immune response in patients with severe COVID-19. Nat Med (2020).

14. Cupovic, J., et al. Central Nervous System Stromal Cells Control Local CD8(+) T Cell Responses during Virus-Induced Neuroinflammation. Immunity 44, 622–633 (2016).

15. Channappanavar, R., et al. Dysregulated Type I Interferon and Inflammatory Monocyte-Macrophage Responses Cause Lethal Pneumonia in SARS-CoV-Infected Mice. Cell Host Microbe 19, 181–193 (2016).

16. Bodnar, R.J., Yates, C.C., Rodgers, M.E., Du, X. & Wells, A. IP-10 induces dissociation of newly formed blood vessels. Journal of cell science 122, 2064–2077 (2009).

17. Yang, Y., et al. Exuberant elevation of IP-10. MCP-3 and IL-1ra during SARS-CoV-2 infection is associated with disease severity and fatal outcome. medRxiv 2002, 2020 (2020).

18. Hueso, L., et al. Upregulation of angiostatic chemokines IP-10/CXCL10 and I-TAC/CXCL11 in human obesity and their implication for adipose tissue angiogenesis. Int J Obes (Lond) 42, 1406–1417 (2018).

19. Bonfante, H.L., et al. CCL2, CXCL8, CXCL9 and CXCL10 serum levels increase with age but are not altered by treatment with hydroxychloroquine in patients with osteoarthritis of the knees. Int J Rheum Dis 20, 1958–1964 (2017).

20. Merad, M. & Martin, J.C. Pathological inflammation in patients with COVID-19: a key role for monocytes and macrophages. Nat Rev Immunol 20, 355–362 (2020).

21. Puthusseri, B., et al. Regulation of p53-mediated changes in the uPA-fibrinolytic system and in lung injury by loss of surfactant protein C expression in alveolar epithelial cells. Am J Physiol Lung Cell Mol Physiol 312, L783–L796 (2017).

22. Ling, G.S., et al. C1q restrains autoimmunity and viral infection by regulating CD8(+) T cell metabolism. Science 360, 558–563 (2018).

23. Diao, B., et al. Reduction and Functional Exhaustion of T Cells in Patients With Coronavirus Disease 2019 (COVID-19). Front Immunol 11, 827 (2020).

24. Salje, H., et al. Estimating the burden of SARS-CoV-2 in France. Science 369, 208–211 (2020).

25. Bosmuller, H., et al. The evolution of pulmonary pathology in fatal COVID-19 disease: an autopsy study with clinical correlation. Virchows Arch, 1–9 (2020).

26. Cervantes-Barragan, L., et al. Control of coronavirus infection through plasmacytoid dendritic-cell-derived type I interferon. Blood 109, 1131–1137 (2007).

27. Kindler, E. & Thiel, V. To sense or not to sense viral RNA--essentials of coronavirus innate immune evasion. Curr Opin Microbiol 20, 69–75 (2014).

28. Cameron, M.J., et al. Interferon-mediated immunopathological events are associated with atypical innate and adaptive immune responses in patients with severe acute respiratory syndrome. J Virol 81, 8692–8706 (2007).

29. Ziegler, C.G.K., et al. SARS-CoV-2 Receptor ACE2 Is an Interferon-Stimulated Gene in Human Airway Epithelial Cells and Is Detected in Specific Cell Subsets across Tissues. Cell 181, 1016–1035 e1019 (2020).

30. Hadjadj, J., et al. Impaired type I interferon activity and exacerbated inflammatory responses in severe Covid-19 patients. MedRxiv (2020).

31. Konopka, K.E., et al. Diffuse Alveolar Damage (DAD) from Coronavirus Disease 2019 Infection is Morphologically Indistinguishable from Other Causes of DAD. Histopathology (2020).

32. Fei, J., et al. Reduction of lymphocyte at early stage elevates severity and death risk of COVID-19 patients: a hospital-based case-cohort study. medRxiv (2020).

33. Zheng, M., et al. Functional exhaustion of antiviral lymphocytes in COVID-19 patients. Cell Mol Immunol 17, 533–535 (2020).

34. Zhao, J., Zhao, J. & Perlman, S. T cell responses are required for protection from clinical disease and for virus clearance in severe acute respiratory syndrome coronavirus-infected mice. J Virol 84, 9318–9325 (2010).

35. Ng, O.W., et al. Memory T cell responses targeting the SARS coronavirus persist up to 11 years post-infection. Vaccine 34, 2008–2014 (2016).

36. Wolfel, R., et al. Virological assessment of hospitalized patients with COVID-2019. Nature 581, 465–469 (2020).

37. Magro, C., et al. Complement associated microvascular injury and thrombosis in the pathogenesis of severe COVID-19 infection: A report of five cases. Transl Res 220, 1–13 (2020).

38. Cugno, M., et al. Complement activation in patients with COVID-19: A novel therapeutic target. J Allergy Clin Immunol 146, 215–217 (2020).

39. Sweeney, R.M. & McAuley, D.F. Acute respiratory distress syndrome. Lancet 388, 2416–2430 (2016).

40. Copin, M.C., et al. Time to consider histologic pattern of lung injury to treat critically ill patients with COVID-19 infection. Intensive Care Med 46, 1124–1126 (2020).

41. Xu, Z., et al. Pathological findings of COVID-19 associated with acute respiratory distress syndrome. Lancet Respir Med 8, 420–422 (2020).

42. Bates, D., Mächler, M., Bolker, B. & Walker, S. Fitting linear mixed-effects models using lme4. arXiv preprint 1406.5823 (2014).

43. Chikina, M., Robinson, J.D. & Clark, N.L. Hundreds of Genes Experienced Convergent Shifts in Selective Pressure in Marine Mammals. Mol Biol Evol 33, 2182–2192 (2016).

44. Team, R.C. R: A language and environment for statistical computing. (2013).

45. Wickham, H. ggplot2: elegant graphics for data analysis, (Springer, 2016).

46. Gu, Z., Gu, L., Eils, R., Schlesner, M. & Brors, B. circlize Implements and enhances circular visualization in R. Bioinformatics 30, 2811–2812 (2014).

47. Gu, Z., Eils, R. & Schlesner, M. Complex heatmaps reveal patterns and correlations in multidimensional genomic data. Bioinformatics 32, 2847–2849 (2016).

48. Horikoshi, M. & Tang, Y. ggfortify: Data visualization tools for statistical analysis results. v0.1.0. URL http://CRAN.R-project.org/package=ggfortify. R package version 0.4 1, 28 (2018).

49. Wickham, H. Reshaping data with the reshape package. Journal of statistical software 21, 1–20 (2007).

50. Kassambara, A. & Mundt, F. Package ‘factoextra’. Extract and visualize the results of multivariate data analyses 76(2017).

